# Projections of human papillomavirus (HPV) vaccination impact on non-cervical cancer outcomes among women in 117 low-income and middle-income countries: a modeling study

**DOI:** 10.1101/2025.06.24.25330224

**Authors:** Aarushi Tuli, Mark Jit, Kaja Abbas, Allison Portnoy

## Abstract

**Introduction:** Human papillomavirus (HPV) is a leading cause of both cervical and non-cervical cancers, including anal, oropharyngeal, vaginal, and vulvar cancers. While most HPV vaccination impact assessments have focused on preventing cervical cancer among women, the broader benefits of vaccination against other HPV-attributable cancers in low- and middle-income countries (LMICs) remain less explored.

**Methods:** We used a static cohort model to assess the potential health impact of bivalent HPV vaccination on HPV-attributable female non-cervical cancers in 117 LMICs from 2030 to 2100. The model incorporated country-specific data on cancer mortality, HPV type distribution, demographic projections, and vaccine coverage. Sensitivity analyses were performed to account for uncertainties in cancer incidence and mortality, HPV type distribution, and cancer stage distribution.

**Results:** Our projections suggest that HPV vaccination could contribute to prevention of approximately 590,000 cases of anal, 880,000 oropharyngeal, 1.27 million vaginal, and 2.18 million vulvar cancers over the analytic period. In total, 3.0 million deaths from these non-cervical cancers among women could be averted by 2100. The African Region is expected to see the largest relative reductions in both cases and deaths, while the European region showed the smallest gains. By the end of the century, 58 countries are projected to reach at least a 25% reduction in anal cancer mortality, compared to just 25 countries for oropharyngeal cancer. These findings reflect substantial regional disparities in both burden and vaccination impact.

**Conclusion:** HPV vaccination holds considerable promise in reducing the burden of non-cervical cancers among women in LMICs, especially in regions with high incidence and limited access to care. Recognizing and harnessing these broader benefits can strengthen the public health case for scaling up vaccine access, implementing region-specific strategies, and investing in equitable healthcare systems to decrease global disparities.

**KEY MESSAGES:** What is Already Known About This Topic?

Health-impact modelling has been used to project the impact of human papillomavirus (HPV) vaccination on prevention of cervical cancer.

HPV is a leading cause of non-cervical cancers, including anal, oropharyngeal, vaginal, and vulvar cancers.

What Does This Study Add?

This study estimated the impact of HPV vaccination on non-cervical female cancers, projecting the largest reductions in the African region.

How Might This Study Affect Research, Practice, or Policy?

In order to improve coverage of HPV vaccination programs, it will be important to estimate additional cancer prevention benefits that can mitigate disease burden in low- and middle-income countries.

## INTRODUCTION

Human papillomavirus (HPV) is a leading cause of cancer-related mortality among women worldwide.^1^ While almost all cervical cancer cases are caused by HPV in women, HPV infection also contributes to several other cancers, including anal, oropharyngeal, vaginal, and vulvar.^2,3^

In settings with robust data, mathematical models have demonstrated the profound health and economic benefits of widespread HPV vaccination.^4^ In settings with limited data, where the burden of HPV-attributable cancer is disproportionately high, the overall impact of HPV vaccination on non-cervical cancers remains underexplored. HPV types 16 and 18 are responsible for the majority of HPV-attributable cancers globally – such as anal, oropharyngeal, vaginal, and vulvar – but are also vaccine-preventable. The bivalent, quadrivalent, and nonavalent HPV vaccines protect against these high-risk HPV types 16 and 18.^4^ Prophylactic HPV vaccination has been shown to provide nearly 100% protection against persistent infection from high-risk HPV types, when administered before exposure to the virus.^5–7^ Vaccination is most effective prior to sexual debut, starting at ages 9–12, as recommended by health organizations worldwide.^4, 8–12^ A bivalent vaccine is designed to protect against two specific virus types; in the case of HPV, the WHO-prequalified bivalent vaccine targets HPV types 16 and 18.^8^ However, while the impact of HPV vaccination on cervical cancer outcomes has been well documented, less attention has been focused on estimating the non-cervical cancer benefits of HPV vaccination especially in LMICs.^9–11,13^

In order to address this gap, we expanded an existing HPV vaccination model to project the estimated health impact of HPV vaccination among women beyond cervical cancer, incorporating anal, oropharyngeal, vaginal, and vulvar cancers. Using this mathematical model, we assessed vaccine-impact projections to estimate the potential health impact of HPV vaccination among vaccinated cohorts of women in 117 LMICs. We estimated the potential health impact in terms of cancer cases and deaths averted among vaccinated cohorts from the time of vaccination until 2100.

## METHODS

### Model Framework

The HPV model employed in this analysis is designed to estimate the health and economic impacts of HPV vaccination in resource-limited settings.^14–16^ This static cohort model compares health outcomes with and without HPV vaccination for female cohorts targeted for vaccination at a specific age (e.g., 9 years) throughout their lifetimes. Instead of fully replicating the natural progression of HPV infection, the model utilizes a simplified decision-tree approach, estimating proportional reduction in non-cervical (anal, oropharyngeal, vaginal, and vulvar) cancer burden across the life course of vaccinated women, incorporating key input data related to demography, HPV-attributable cancer incidence, HPV type distribution, and probability of death. These assumptions are adjusted in probabilistic sensitivity analyses (PSA) to account for uncertainties in model parameters (Figure S1 and Appendix Table S1).

The model framework provides a structured approach to estimate the impact of female HPV vaccination on reducing lifetime cancer risks associated with HPV. It compares two scenarios: no vaccination (resulting in no reduction in cancer risk) and vaccination (resulting in a reduction in cancer cases). Key model inputs include vaccine efficacy, coverage, country-specific HPV-16/18 type distribution, and cancer incidence for anal, oropharyngeal, vaginal, and vulvar cancers.

The incidence is applied annually, and cases and deaths are cumulatively calculated in the vaccination and no-vaccination scenarios to estimate cases and deaths averted by vaccination. The model estimates vaccination impact in terms of reductions in age-dependent cervical cancer incidence and mortality in direct proportion to vaccine efficacy, vaccine coverage, and HPV type distribution. Age-specific population size estimates (in 1-year intervals) were sourced from the United Nations World Population Prospects 2019 revision, while age-specific life expectancy (in five-year intervals) was obtained from 2019 World Health Organization (WHO) life tables and held constant across the time horizon of the analysis.^17,18^

Additional inputs incorporate country-specific population projections reflecting age structure and background mortality, local age-specific life expectancy, and the probability of cancer death. These inputs enable calculations of the number of cancer cases and deaths averted due to vaccination (Figure S1). The parameters and model assumptions are summarized in Table 1.

**Table 1.** Parameters/model assumptions

| **Input** | **Value/Assumption/Formula** | **Source** | **Detailed Information** |
| --- | --- | --- | --- |
| **Vaccination coverage** | Country-specific vaccination coverage and year of HPV vaccine introduction | [18,19, 24–28] | Methods |
| **HPV type distribution** | Relative contributions of HPV types 16 and 18; in case of missing data, regional data applied to corresponding countries | [21–23] | Appendix Tables S3–S6 |
| **Vaccine efficacy** | Lifelong 100% efficacy against HPV 16/18 infections | [29–32] | Methods |
| **Age-specific cancer incidence rates** | GLOBOCAN database; income category defined by 2022–2033 World Bank income classification; estimates based on regional and income category averages for countries without data available on GLOBOCAN | [19,20] | Appendix Table S2 |
| **Cause-specific probability of death** | Mortality rates (per 100,000 women per year) used to estimate probability of death | [20–21] | Appendix Table S8 |

### Age-Specific Cancer Incidence Rates

Estimates of age-specific cancer (anal, oropharyngeal, vaginal, and vulvar) incidence rates were sourced from the GLOBOCAN database, provided by the Global Cancer Observatory of the International Agency for Research on Cancer (IARC) at the World Health Organization (WHO).^19^ The income category for each country was defined by the 2022–2023 World Bank income classification.^20^ For countries without estimates available on GLOBOCAN, estimates were made based on regional and income category averages as detailed in Appendix Table S2.

### HPV Type Distribution in Non-Cervical Cancer

The relative contributions of HPV types 16 and 18 to each non-cervical cancer were based on de Sanjosé, et al. (2019).^21^ For oropharyngeal cancer type distribution in African Region (AFR), additional assumptions were based on de Martel, et al. (2017) and Castellsagué, et al. (2016).^21,22^ In cases of missing data, regional data from Europe, the Region of the Americas (AMR), Asia, and Oceania were applied to corresponding countries. For the Eastern Mediterranean Region (EMR), values were assigned based on the country’s continent. Appendix Tables S3–S6 define the full list of type distribution assumptions by country.

For endpoints other than oropharyngeal cancer in AFR, data from Europe and the AMR were applied to countries within these regions for the type distribution.^21^Asia was used for countries in both Southeast Asia Region (SEAR) and Western Pacific Region (WPR), depending on their geographic location, with Oceania used for the remaining WPR countries. For EMR, values were assigned based on the continent of the country.^21^ For anal cancer, the type distribution estimated by de Sanjosé, et al., was not female-specific, so it was assumed that the type distribution for all (male and female) anal cancers applied to females.^21^ For oropharyngeal cancer in AFR, the mean estimate of HPV attribution was set at 13% for all AFR countries, per de Martel, et al. (2017).^22^ Although this estimate accounts for all HPV types rather than being HPV-16/18 specific, HPV-16 is predominantly responsible for HPV-attributable oropharyngeal cancers globally.^22^ The lower bound was assumed to be zero per de Sanjosé et al. (2019), and the upper bound was set to 25% per Castellsagué et al. (2016), based on HPV-DNA positivity and either E6*I mRNA+ or p16^INK4a^+ biomarkers in oropharyngeal cancers.^23^ For type distribution assumptions, we based our assumptions on the advanced molecular and immunohistochemical assays for detecting and characterizing HPV in cancer samples as per one study.^21^ Key methods included SPF-10 PCR with DNA enzyme immunoassay (DEIA) for broad-spectrum HPV DNA detection and the LiPA25 reverse hybridization assay to genotype 25 HPV types. To confirm viral activity, the E6*I mRNA assay targeted transcriptional biomarkers, while p16INK4a immunohistochemistry assessed cellular markers of HPV-driven transformation. These comprehensive techniques ensured accurate identification and distribution analysis of HPV types across various cancer cases.^21^ We assume that estimates for AFR countries do not go beyond the global upper bound, keeping in mind that more recent data are likely to use these improved methods.

### Coverage

We assumed country-specific vaccination coverage and year of HPV vaccine introduction based on historical estimates from WHO and UNICEF Estimates of National Immunization Coverage (WUENIC) for routine vaccination and the WHO immunization repository for campaign vaccination for all years up to 2022.^24,25^ From 2023–2030, we assumed projected coverage informed by the WHO immunization repository, WHO Immunization Agenda 2030 targets, and expert consultation.^26^ Beyond 2030, we projected that routine coverage of HPV vaccine would increase at a rate of 1 percentage point annually through 2100, with an upper limit of 95%. These coverage assumptions were developed by the Vaccine Impact Modelling Consortium per previously published analyses.^27,28^ The model assumes lifelong 100% efficacy of the vaccine against HPV-16/18 infections.^29–32^

### Cause-Specific Probability of Death

To estimate cancer mortality, the model first assumed country-specific distributions of cancer stages for incident cancers across the time horizon of the analysis (Appendix Table S7). Mortality rates (per 100,000 women per year) attributable to anal, oropharyngeal, vaginal, and vulvar cancers were used to estimate probability of death from the HPV Stats database.^33^ Average values from countries within the same region and income category were used when data were missing from the HPV Stats website. Adjustments were made for countries and outcomes where data were unavailable (Appendix Table S8). Background mortality was indirectly incorporated through the use of population projections from the United Nations World Population Prospects that incorporate births and deaths over time.^34^

### Sensitivity Analysis

Three key parameters were identified for PSA: HPV-16/18 type distribution, age-specific cancer incidence (for each cancer type), and probability of death. Each parameter was assigned a beta-PERT distribution for probabilistic sampling, with the bounds determined by (1) standard deviations across empirical data for type distribution; (2) confidence intervals estimated from cancer cases in GLOBOCAN;^19^ and (3) standard deviations across estimated probabilities for probability of death. Two hundred independent parameter sets were drawn, and 95% uncertainty intervals (UIs) were extracted from the 2.5 and 97.5 percentiles of the sample results obtained.

## RESULTS

We estimated that 590,000 (95% uncertainty interval: 560,000–630,000) anal, 880,000 (820,000–950,000) oropharyngeal, 1.27 (1.20–1.35) million vaginal, and 2.18 (2.09–2.29) million vulvar cancer cases could be averted by bivalent HPV vaccination across 117 LMICs in 2030–2100 (Table 2). We also estimated that 360,000 (340,000–390,000) anal, 550,000 (510,000–600,000) oropharyngeal, 800,000 (760,000–860,000) vaginal, and 1.30 (1.24–1.36) million vulvar cancer deaths could be averted by bivalent HPV vaccination across 117 LMICs in 2030–2100.

**Table 2:** Regional impact of HPV vaccination on cancer outcomes (2030–2100).

| Region | Cases Averted (millions) |  |  |  |  |
| --- | --- | --- | --- | --- | --- |
|  | Anal Cancer | Oropharyngeal Cancer | Vaginal Cancer | Vulvar Cancer | Total |
| **AFR** | 0.33 (0.30–0.36) | 0.38 (0.35–0.41) | 0.54 (0.51–0.56) | 1.07 (1.03–1.13) | 2.32 (2.19–2.45) |
| **AMR** | 0.03 (0.03–0.03) | 0.04 (0.04–0.05) | 0.06 (0.05–0.06) | 0.09 (0.08–0.09) | 0.22 (0.20–0.23) |
| **EMR** | 0.06 (0.05–0.06) | 0.09 (0.08–0.10) | 0.13 (0.12–0.14) | 0.22 (0.20–0.23) | 0.49 (0.45–0.53) |
| **EUR** | 0.02 (0.02–0.02) | 0.03 (0.02–0.03) | 0.03 (0.03–0.03) | 0.05 (0.04–0.05) | 0.12 (0.11–0.13) |
| **SEAR** | 0.12 (0.10–0.14) | 0.28 (0.23–0.34) | 0.44 (0.37–0.51) | 0.63 (0.57–0.72) | 1.47 (1.26–1.71) |
| **WPR** | 0.04 (0.03–0.04) | 0.06 (0.05–0.06) | 0.08 (0.07–0.09) | 0.13 (0.12–0.15) | 0.31 (0.27–0.34) |
| **Total** | 0.59 (0.56–0.63) | 0.88 (0.82–0.95) | 1.27 (1.20–1.45) | 2.18 (2.09–2.29) | 4.92 (4.66–5.21) |

| Region | Deaths Averted (millions) |  |  |  |  |
| --- | --- | --- | --- | --- | --- |
|  | Anal Cancer | Oropharyngeal Cancer | Vaginal Cancer | Vulvar Cancer | Total |
| **AFR** | 0.19 (0.17–0.20) | 0.23 (0.21–0.24) | 0.33 (0.31–0.35) | 0.60 (0.57–0.63) | 1.34 (1.26–1.43) |
| **AMR** | 0.02 (0.02–0.02) | 0.03 (0.02–0.03) | 0.04 (0.03–0.04) | 0.05 (0.05–0.06) | 0.13 (0.12–0.14) |
| **EMR** | 0.04 (0.04–0.05) | 0.07 (0.06–0.07) | 0.09 (0.09–0.10) | 0.15 (0.14–0.16) | 0.35 (0.32–0.38) |
| **EUR** | 0.01 (0.01–0.01) | 0.02 (0.01–0.02) | 0.02 (0.02–0.02) | 0.03 (0.02–0.03) | 0.07 (0.06–0.08) |
| **SEAR** | 0.08 (0.07–0.10) | 0.18 (0.15–0.22) | 0.27 (0.23–0.32) | 0.39 (0.35–0.45) | 0.92 (0.79–1.08) |
| **WPR** | 0.02 (0.02–0.03) | 0.04 (0.03–0.04) | 0.05 (0.05–0.06) | 0.09 (0.08–0.10) | 0.21 (0.18–0.23) |
| **Total** | 0.36 (0.34–0.39) | 0.55 (0.51–0.60) | 0.80 (0.76–0.86) | 1.30 (1.24–1.36) | 3.02 (2.85–3.20) |

\* Figures are rounded to the nearest ten thousand.
Note: WHO refers to the World Health Organization. Regional abbreviations are as follows: AFR (African Region), AMR (Region of the Americas), EMR (Eastern Mediterranean Region), EUR (European Region), SEAR (South-East Asia Region), and WPR (Western Pacific Region).

The total number of cases and deaths averted due to HPV vaccination was variable across World Health Organization (WHO) regions, reflecting reflecting differences in demography, epidemiological and healthcare contexts. The African Region (AFR) had the highest percentage reduction overall for both cases (42.8% for anal, 30.8% for oropharyngeal, 32.8% for vaginal, 44.2% for vulvar cancer) and deaths (13.8% for anal, 16.8% for oropharyngeal, 24.7% for vaginal, and 44.7% for vulvar cancer) followed closely by SEAR and EMR. WPR had the lowest percentage reductions for cases (11.2% for anal, 8.8% for oropharyngeal, 9.4% for vaginal and 10.3% for vulvar cancer) and for deaths (12% for anal, 18.3% for oropharyngeal, 26.9% for vaginal and 42.8% for vulvar cancer).

The estimates also revealed regional differences in the impact of HPV vaccination on specific cancer types. Notably, AFR showed high numbers of anal (330,000 [300,000–360,000]) and vulvar (1.07 million [1.03–1.13]) cancers averted. The impact on oropharyngeal and vaginal cancers was substantial across all regions.

The bar chart presented in Figure 1 illustrates the comparative analysis of four different cancer types across the six WHO regions. For all the regions, vulvar cancer was estimated to have the greatest reduction in deaths due to HPV vaccination, whereas AMR and EUR showed greatest impact on anal cancer.

**Figure 1:**
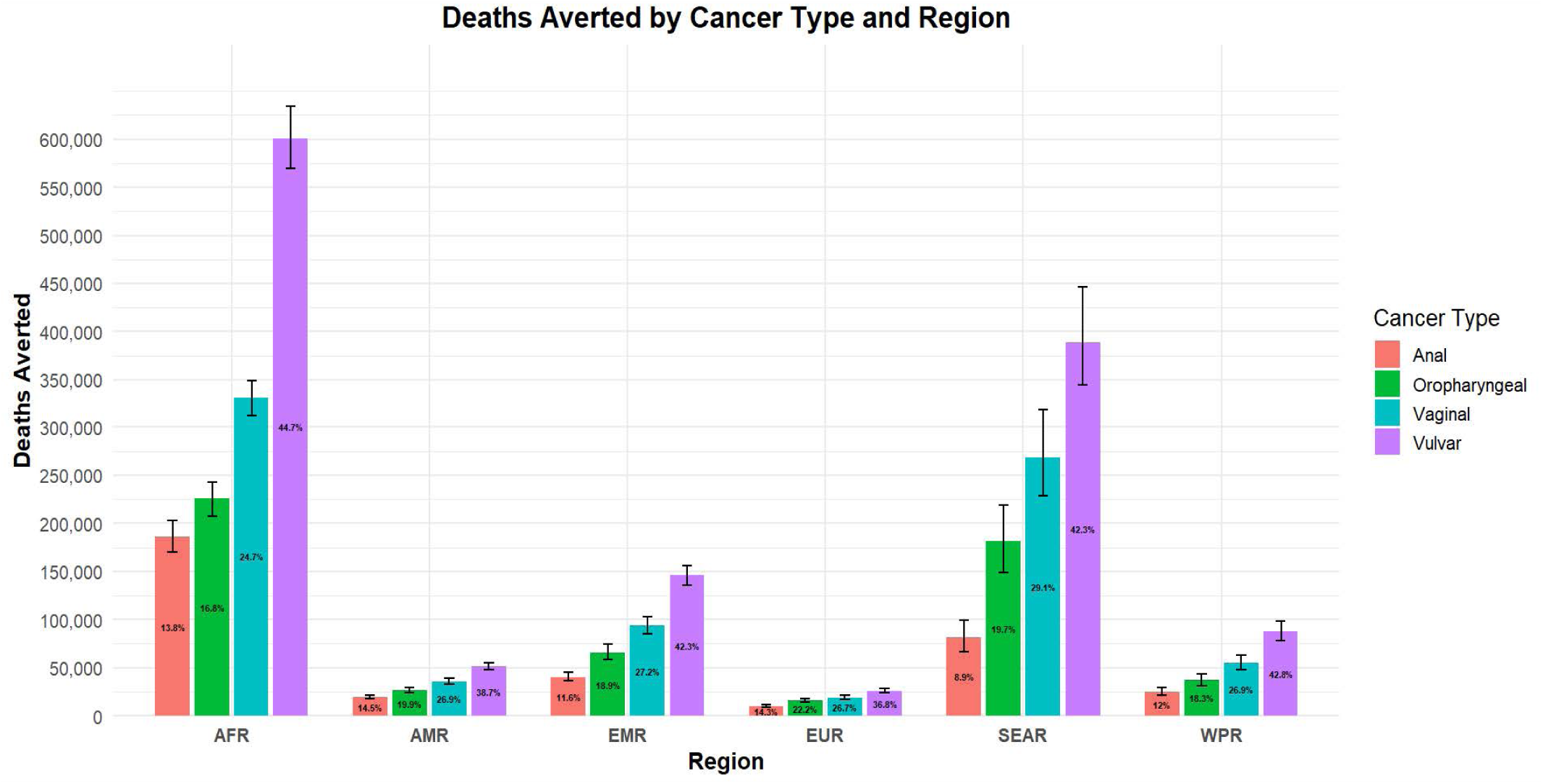
Reduction in cancer deaths by cancer type.

The bar graph in Appendix Figure S2 illustrates the top 10 countries with the highest percentage of deaths averted by HPV vaccination for each cancer type. The countries represented include India, Nigeria, Ethiopia, Pakistan, Democratic Republic of the Congo, China, United Republic of Tanzania, Indonesia, Uganda, and Kenya.

Figure 2 illustrates the geographic distribution of the impact of HPV vaccination on reducing non-cervical deaths among the 117 LMICs included in the analysis. The map is color-coded, highlighting variations in mortality reduction due to the vaccine’s implementation in various countries. The majority of countries in Sub-Saharan Africa showed a comparatively higher percentage of deaths averted due to HPV vaccination, around 30% or greater. Countries in the AMR had 12–17%, EMR had 22–29%, EUR had 14–26%, and SEAR had 14–20% reduction in deaths for all cancers respectively. Countries in the WPR had less than 10% reduction in deaths for all cancers.

**Figure 2:**
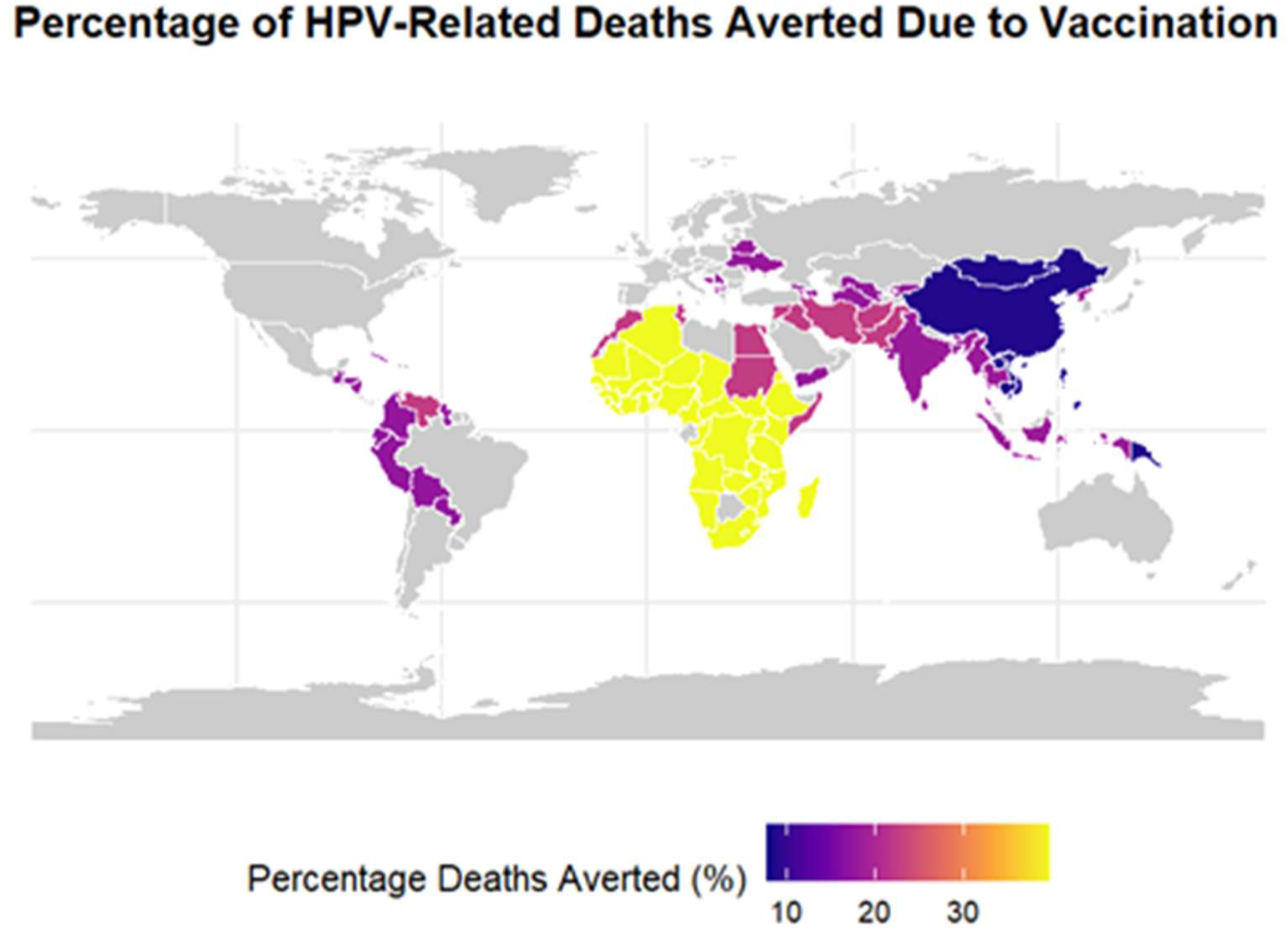
Geographic distribution of reduction in anal, oropharyngeal, vaginal, and vulvar cancer deaths due to HPV vaccination.

Figure 3 depicts the cumulative number of countries reaching 25% reduction in deaths over time due to the four cancer types. For anal cancer, 58 countries achieved a 25% reduction in deaths by the year 2100. For vulvar cancer, 47 countries achieved a 25% reduction in deaths slightly before the year 2100. For vaginal cancer, 41 countries achieved a 25% reduction in deaths by the year 2100. However, for oropharyngeal cancer, only 25 countries achieved a 25% reduction in cancer by the year 2100. For all the remaining countries, a 25% reduction in deaths was not achieved before the year 2100.

**Figure 3:**
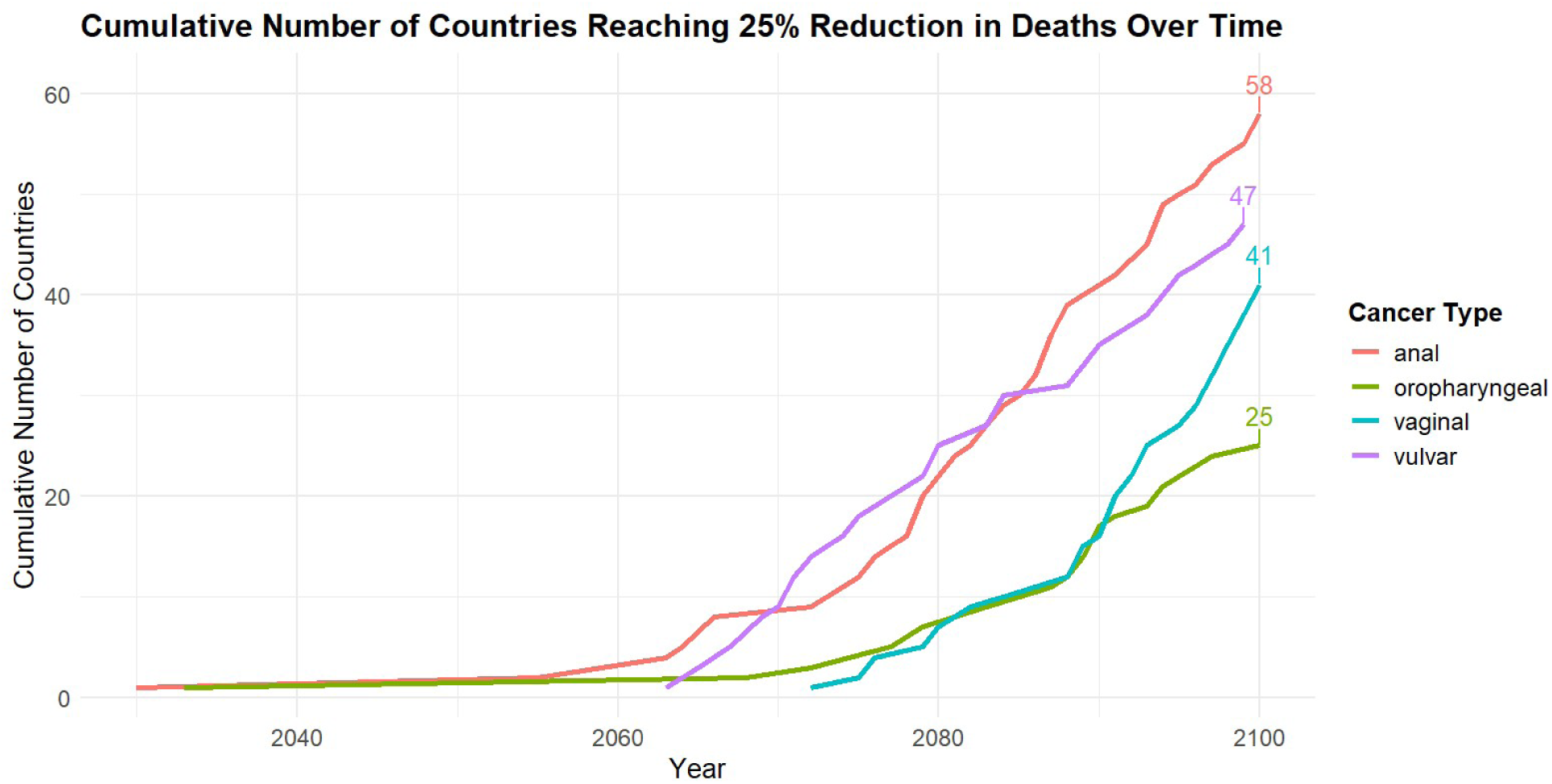
A line graph showing the cumulative number of countries reaching 25% reduction in deaths due to HPV attributable anal, oropharyngeal, vaginal and vulvar cancers over time for the time period 2030 to 2100. *Note: Numeric labels reflect the number of countries reaching 25% reduction before the year 2100.

## DISCUSSION

We conducted an assessment of HPV vaccination’s broader impact in averting the burden of female cancers beyond cervical cancer from 2030 to 2100 in 117 LMICs. Over this timeframe, vaccination is projected to avert 590,000 (560,000–630,000) anal, and 880,000 (820,000–950,000) oropharyngeal, 1.27 (1.20–1.35) million vaginal, and 2.18 (2.09–2.29) million vulvar cancer cases, along with 360,000 (340,000–390,000) anal, 550,000 (510,000–600,000) oropharyngeal, 800,000 (760,000–860,000) vaginal, and 1.30 (1.24–1.36) million vulvar cancer deaths.

While cervical cancer has been the primary focus of HPV vaccination efforts, averting an estimated 45.8 million deaths (a 64% reduction) across 78 LMICs from 2020 to 2120, our findings highlight the substantial, yet often overlooked, impact on non-cervical cancers. In 117 LMICs, HPV vaccination is projected to avert 3.02 million deaths from non-cervical cancers by 2100. By 2100, 58 countries are expected to achieve at least a 25% reduction in anal cancer deaths, comapred with 47 countries achieving this reduction in vulvar, 41 in vaginal, and 25 in oropharyngeal cancers. These results highlight the broader protective potential of HPV vaccination and the importance of extending research and policy attention beyond cervical cancer to fully realize its benefits across all HPV-related cancers.^10^

The most substantial reductions were estimated in AFR, followed by SEAR and EMR, while EUR had the lowest projected impact. Specific cancer type impacts vary, with anal and vulvar cancer showing significant reductions in AFR and SEAR. This study’s added value lies in its comprehensive assessment of HPV vaccination’s broader impact in averting female cancers beyond cervical cancer, emphasizing regional differences and highlighting the potential of vaccination programs to significantly reduce cancer burdens across diverse settings. The analysis highlights the profound public health impact of HPV vaccination in preventing multiple HPV-related cancers, highlighting the importance of targeted vaccination programs to address regional disparities in healthcare access and vaccine coverage.

When looking at the averted cases and deaths of anal, oropharyngeal, vaginal, and vulvar cancers, the robust impact projected in AFR could be attributed to the higher burden and lower level of vaccine adoption, highlighting disparities in vaccine access. While the impact remains significant in AFR and SEAR, there may be gaps due to lower projected HPV vaccination coverage, despite high burden particularly in AFR.^35,36^ The substantial impact on vaginal and vulvar cancers across all regions indicates the broad-spectrum benefits of HPV vaccination in preventing HPV-related female cancers beyond cervical cancer. Oropharyngeal cancer is now the most common HPV associated cancer in high-income countries.^34^ In over 90% of cases, HPV-related oropharyngeal cancer is caused by oral infection with HPV-16, which is also the primary type involved in the vast majority (90%) of these cases.^37^ It is also worth noting the rising incidence of HPV-related anal cancer driven largely by HPV16 in high-income countries like Australia, Canada, and the U.S., where nearly all cases are HPV-positive.^38^ This trend may eventually emerge in LMICs as healthcare systems evolve, reinforcing the need for timely HPV vaccine rollout and scale-up.

The variation in the results across different regions indicates that one-size-fits-all policies may not be effective. Tailored approaches that consider regional specificities and needs are essential. The substantial projected impact in AFR highlights the importance of scale-up and targeted outreach in high-need settings. The variation in deaths averted by cancer type across regions highlights potential disparities that need to be addressed. Policymakers should focus on promoting equity by ensuring that all regions have access to the necessary resources and support to improve their performance. The large reductions in oropharyngeal, vaginal, and vulvar cancers across all regions demonstrate the broad benefits of HPV vaccination beyond cervical cancer prevention and can inform equitable resource allocation by directing investments toward regions and populations with the greatest potential health gains. The broader impact across multiple HPV-attributable cancers highlights the importance of health system decision-making that advances equity by allocating resources to address the full spectrum of HPV-related cancers. For Gavi, the Vaccine Alliance, capturing these additional cancer outcomes is also important to more comprehensively reflect the lives saved and the broader health impact achieved through Gavi-supported HPV vaccination programs. Targeted interventions should aim to reduce disparities and promote balanced development across all regions. For example, these may include catch-up campaigns for older adolescents or young adults, and targeted outreach to underserved or hard-to-reach populations, tailored to each region’s specific challenges and resource constraints. The substantial benefits highlight the critical need for continued investment in vaccination programs and access to healthcare services. In settings with restricted access to facility-based care, mainly in parts of AFR and SEAR, school-based vaccination programs can provide an effective platform to reach adolescents. Community-based or outreach distribution may be essential in parts with geographically spread populations. A potential health systems reform could involve integrating HPV vaccination into existing adolescent health or antenatal care programs to improve coverage, streamline delivery, and promote more equitable access.^39^

This study used a static cohort model and did not account for indirect protection (herd immunity from female vaccination or the additional benefits to females from male vaccination). Several modelling studies have examined differences between static cohort models and dynamic transmission models of HPV vaccination. Dynamic models capture herd immunity, whereas static models do not, but when routine immunization achieves high coverage in young girls (e.g., ≥ 80 % in ages 9–14 years), projected outcomes from static models are similar to dynamic models because herd effects are near maximal.^40, 41^

We did not include other HPV-attributable outcomes in females, such as genital warts, which would further increase the estimated benefits of vaccination. We also excluded the additional benefits of gender-neutral vaccination in preventing cancers among females, as well as its estimated impact on cancers among males. We did not examine cancer screening programs in this analysis and assumed that any ongoing screening programs did not change as HPV vaccination introduction and delivery changed. We assumed constant age-specific incidence rates, stage distribution, HPV type distribution, and mortality rates. In practice, these parameters may change as vaccination coverage increases and screening programs expand, particularly in LMICs, potentially shifting diagnoses to earlier stages and reducing mortality. Our estimates may overestimate the impact of HPV vaccination on anal, oropharyngeal, vaginal, and vulvar cancer by holding these assumptions constant but may underestimate impact due to excluding indirect effects.

This study made several assumptions due to the lack of availability of specific data, and these data gaps add uncertainty to the inferences on HPV vaccination impact on anal, oropharyngeal, vaginal, and vulvar cancers. Additionally, for countries lacking GLOBOCAN estimates, incidence rates were inferred from regional and income category proxies, which may not fully capture the local epidemiological context.

The study relied on regional estimates for HPV type distribution, which might not reflect the local diversity in circulating HPV types. For oropharyngeal cancer in AFR, the type distribution was generalized from broader estimates, introducing potential inaccuracies, particularly given the global variability in HPV-16 prevalence. In the absence of country-specific mortality estimates, average mortality rates from similar regions and income categories were used. Also, the use of non-female-specific type distribution for anal cancer may overestimate incidence in females.^21^

These assumptions, while necessary for filling data gaps, introduced limitations that should be considered when interpreting the findings. In particular, extrapolating incidence and other epidemiological inputs from countries with available data to those without may introduce bias if countries lacking data differ systematically from those used as the basis for extrapolation. This limitation is not unique to our study but reflects a broader challenge across HPV modelling studies, both static and dynamic, as it stems from constraints in the underlying epidemiological datasets rather than the modelling framework itself. These findings should be interpreted primarily as qualitative indications of the relative magnitude and regional distribution of health impact rather than precise quantitative estimates for any individual country, particularly given that key inputs were unavailable for some settings and therefore required extrapolation from available data. Future research to expand country-level primary data collection will be essential to generate more precise estimates and support evidence-based policy decisions.

Our study contributes to the growing evidence supporting the broader benefits of HPV vaccination, reinforcing its role as a crucial tool in reducing global cancer burden. To maximize these benefits, international cooperation, increased investment, and targeted policy interventions are essential to ensure equitable access to HPV vaccination and to harness its full potential to improve global health outcomes. Overall, the substantial impact of HPV vaccination on preventing cancer outcomes across diverse regions highlights the global benefits of this intervention. Continued efforts to improve vaccination coverage and healthcare access will be crucial in reducing the HPV-attributable cancer burden worldwide and achieving equitable health outcomes.

While cervical cancer remains the primary target of HPV vaccination programs, accounting for the prevention of anal, oropharyngeal, vaginal, and vulvar cancers highlights a more comprehensive public health impact, increasing the number of cases and deaths averted across diverse regions and cancer types. This expanded scope not only highlights the vaccine’s potential to reduce the total HPV-related cancer burden but also enhances its cost-effectiveness by capturing additional health gains that might otherwise be overlooked. Incorporating these outcomes provides a stronger economic and health rationale for sustained and expanded vaccine implementation, especially in low- and middle-income countries where the burden of HPV-related cancers is high. Ultimately, this holistic perspective supports more robust policy decisions, encourages greater investment, and promotes equitable access to HPV vaccination worldwide.

## CONTRIBUTORS

AT and AP conceptualized and designed the study. AT conducted the analysis with support from AP. AT drafted the initial manuscript and approved the final manuscript as submitted. MJ, KA, and AP critically reviewed the analysis, reviewed and revised the manuscript, and approved the final manuscript as submitted. All authors read and approved the final manuscript. AP accepts full responsibility for the finished work and/or the conduct of the study, had access to the data, and controlled the decision to publish. AP acted as guarantor.

## ETHICS APPROVAL

Not applicable.

## DATA SHARING STATEMENT

Data are available upon reasonable request. The datasets generated during and/or analyzed during the current study are available from the corresponding author on reasonable request.

## CLINICAL TRIAL REGISTRATION

Not applicable.

## AI USE STATEMENT

Artificial intelligence (AI) tools were not used in the preparation of this manuscript.

## ACKNOWLEDGEMENTS

We are grateful for the support of the Vaccine Impact Modelling Consortium for funding this research. We thank the Boston University School of Public Health Career and Practicum Office for guidance and oversight of this research project for Aarushi Tuli’s practicum experience. KA is supported by the Japan Agency for Medical Research and Development (JP223fa627004).

## OTHER STATEMENTS

### Patient and Public Involvement

Patients and/or the public were not involved in the design, or conduct, or reporting, or dissemination plans of this research.

### Patient Consent for Publication

Not applicable.

