## Supplementary Material Appendix for "Projections of human papillomavirus (HPV) vaccination impact on non-cervical cancer outcomes among women in 117 low-income and middle-income countries: a modeling study"

| Parameter | Description of Assumption |
|---|---|
| Cohort Structure | The assumption is that girls fully immunized at a target age and tracked through their lifetimes. The model assumes full vaccination and lifelong immunity. |
| HPV Type Distribution | HPV-16/18 types are attributed to cervical cancer cases. |
| Vaccine Cross-Protection | There is no cross-protection assumed against non-vaccine HPV types. |
| Probabilistic Sensitivity Analysis (PSA) | Parameters are sampled using b-PERT distribution with bounds based on empirical data and assumptions. There are 200 iterations per country, with +/- 10% variation. |

**Table S1:** Summary of key model assumptions.

| Country | Region, Income Category | Matched Country | Cancer Type |
|---|---|---|---|
| Grenada | AMR, umi | Guatemala | Oropharyngeal |
| Kiribati | WPR, lmi | Cambodia | Anal, oropharyngeal, vaginal, and vulvar |
| Marshall Islands | WPR, umi | China | Oropharyngeal, vaginal, and vulvar |
| Tonga | WPR, umi | China | Anal, oropharyngeal, vaginal, and vulvar |
| Tuvalu | WPR, umi | China | Anal, oropharyngeal, vaginal, and vulvar |
| St Vincent & the Grenadines | AMR, umi | Ecuador | Anal, oropharyngeal, vaginal, and vulvar |
| Grenada | AMR, umi | Colombia | Anal, vaginal, vulvar |
| Palestine | EMR, lmi | Egypt | Anal, oropharyngeal, vaginal, and vulvar |
| Kosovo | EUR | Serbia (neighboring country) | Anal, oropharyngeal, vaginal, and vulvar |

**Table S2:** Region and income-level matched country assumptions for countries with missing data for age-specific incidence rate by cancer type.

Note: AMR= Region of the Americas; EMR= Eastern Mediterranean Region; EUR= European Region; lmi= lower-middle income; umi= upper-middle income; WPR= Western Pacific Region.

| Country Name | Region | Income Level | Type distribution mean | Lower Bound | Upper Bound | Source |
|---|---|---|---|---|---|---|
| Afghanistan | EMR | low | 0.9 | 0.85 | 0.95 | 16 (SEAR average) * |
| Angola | AFR | lmi | 0.78 | 0.73 | 0.83 | 16 |
| Albania | EUR | umi | 0.91 | 0.86 | 0.96 | 16 |
| Armenia | EUR | umi | 0.91 | 0.86 | 0.96 | 16 |
| Azerbaijan | EUR | umi | 0.91 | 0.86 | 0.96 | 16 |
| Burundi | AFR | low | 0.78 | 0.73 | 0.83 | 16 |
| Benin | AFR | lmi | 0.78 | 0.73 | 0.83 | 16 |
| Burkina Faso | AFR | low | 0.78 | 0.73 | 0.83 | 16 |
| Bangladesh | SEAR | lmi | 0.9 | 0.85 | 0.95 | 16 |
| Bosnia and Herzegovina | EUR | umi | 0.91 | 0.86 | 0.96 | 16 |
| Belarus | EUR | umi | 0.91 | 0.86 | 0.96 | 16 |
| Belize | AMR | umi | 0.84 | 0.79 | 0.89 | 16 |
| Bolivia, Plurinational State of | AMR | lmi | 0.84 | 0.79 | 0.89 | 16 |
| Bhutan | SEAR | lmi | 0.9 | 0.85 | 0.95 | 16 |
| Central African Republic | AFR | low | 0.78 | 0.73 | 0.83 | 16 |
| China (Asia) | WPR | umi | 0.9 | 0.85 | 0.95 | 16 |
| Cote d'Ivoire | AFR | lmi | 0.78 | 0.73 | 0.83 | 16 |
| Cameroon | AFR | lmi | 0.78 | 0.73 | 0.83 | 16 |
| Congo, the Democratic Republic of the | AFR | low | 0.78 | 0.73 | 0.83 | 16 |
| Congo | AFR | lmi | 0.78 | 0.73 | 0.83 | 16 |
| Colombia | AMR | umi | 0.84 | 0.79 | 0.89 | 16 |
| Comoros | AFR | lmi | 0.78 | 0.73 | 0.83 | 16 |
| Cabo Verde | AFR | lmi | 0.78 | 0.73 | 0.83 | 16 |
| Cuba | AMR | umi | 0.84 | 0.79 | 0.89 | 16 |
| Djibouti | EMR | lmi | 0.78 | 0.73 | 0.83 | 16 (AFR average) * |
| Dominica | AMR | umi | 0.84 | 0.79 | 0.89 | 16 |
| Algeria | AFR | lmi | 0.78 | 0.73 | 0.83 | 16 |
| Ecuador | AMR | umi | 0.84 | 0.79 | 0.89 | 16 |
| Egypt | EMR | lmi | 0.78 | 0.73 | 0.83 | 16 (AFR average) * |
| Eritrea | AFR | low | 0.78 | 0.73 | 0.83 | 16 |
| Ethiopia | AFR | low | 0.78 | 0.73 | 0.83 | 16 |
| Fiji | WPR | umi | 0.9 | 0.85 | 0.95 | 16 |

| | | | | | | |
|---|---|---|---|---|---|---|
| Micronesia, Federated States of | WPR | lmi | 0.9 | 0.85 | 0.95 | 16 |
| Georgia | EUR | umi | 0.91 | 0.86 | 0.96 | 16 |
| Ghana | AFR | lmi | 0.78 | 0.73 | 0.83 | 16 |
| Guinea | AFR | lmi | 0.78 | 0.73 | 0.83 | 16 |
| Gambia | AFR | low | 0.78 | 0.73 | 0.83 | 16 |
| Guinea-Bissau | AFR | low | 0.78 | 0.73 | 0.83 | 16 |
| Grenada | AMR | umi | 0.84 | 0.79 | 0.89 | 16 |
| Guatemala | AMR | umi | 0.84 | 0.79 | 0.89 | 16 |
| Guyana | AMR | | 0.84 | 0.79 | 0.89 | 16 |
| Honduras | AMR | lmi | 0.84 | 0.79 | 0.89 | 16 |
| Haiti | AMR | lmi | 0.84 | 0.79 | 0.89 | 16 |
| Indonesia | SEAR | umi | 0.9 | 0.85 | 0.95 | 16 |
| India | SEAR | lmi | 0.9 | 0.85 | 0.95 | 16 |
| Iran, Islamic Republic of | EMR | lmi | 0.9 | 0.85 | 0.95 | 16 (SEAR average) * |
| Iraq | EMR | umi | 0.9 | 0.85 | 0.95 | 16 (SEAR average) * |
| Jamaica | AMR | umi | 0.84 | 0.79 | 0.89 | 16 |
| Jordan | EMR | lmi | 0.9 | 0.85 | 0.95 | 16 |
| Kenya | AFR | lmi | 0.78 | 0.73 | 0.83 | 16 |
| Kyrgyzstan | EUR | | 0.91 | 0.86 | 0.96 | 16 |
| Cambodia | WPR | lmi | 0.9 | 0.85 | 0.95 | 16 |
| Kiribati | WPR | lmi | 0.9 | 0.85 | 0.95 | 16 |
| Lao People's Democratic Republic | WPR | lmi | 0.9 | 0.85 | 0.95 | 16 |
| Liberia | AFR | low | 0.78 | 0.73 | 0.83 | 16 |
| St. Lucia | AMR | umi | 0.84 | 0.79 | 0.89 | 16 |
| Sri Lanka | SEAR | lmi | 0.9 | 0.85 | 0.95 | 16 |
| Lesotho | AFR | lmi | 0.78 | 0.73 | 0.83 | 16 |
| Morocco | EMR | lmi | 0.78 | 0.73 | 0.83 | 16 (AFR average) * |
| Moldova, Republic of | EUR | umi | 0.91 | 0.86 | 0.96 | 16 |
| Madagascar | AFR | low | 0.78 | 0.73 | 0.83 | 16 |
| Maldives | SEAR | umi | 0.9 | 0.85 | 0.95 | 16 |
| Marshall Islands | WPR | umi | 0.9 | 0.85 | 0.95 | 16 |
| Macedonia, the former Yugoslav Republic of | EUR | | 0.91 | 0.86 | 0.96 | 16 |
| Mali | AFR | low | 0.78 | 0.73 | 0.83 | 16 |

| | | | | | | |
|---|---|---|---|---|---|---|
| Myanmar | SEAR | lmi | 0.9 | 0.85 | 0.95 | 16 |
| Mongolia | WPR | lmi | 0.9 | 0.85 | 0.95 | 16 |
| Mozambique | AFR | low | 0.78 | 0.73 | 0.83 | 16 |
| Mauritania | AFR | lmi | 0.78 | 0.73 | 0.83 | 16 |
| Malawi | AFR | low | 0.78 | 0.73 | 0.83 | 16 |
| Namibia | AFR | umi | 0.78 | 0.73 | 0.83 | 16 |
| Niger | AFR | low | 0.78 | 0.73 | 0.83 | 16 |
| Nigeria | AFR | lmi | 0.78 | 0.73 | 0.83 | 16 |
| Nicaragua | AMR | lmi | 0.84 | 0.79 | 0.89 | 16 |
| Nepal | SEAR | lmi | 0.9 | 0.85 | 0.95 | 16 |
| Pakistan | EMR | lmi | 0.9 | 0.85 | 0.95 | 16 (SEAR average) * |
| Peru | AMR | umi | 0.84 | 0.79 | 0.89 | 16 |
| Philippines (Asia) | WPR | lmi | 0.9 | 0.85 | 0.95 | 16 |
| Papua New Guinea | WPR | lmi | 0.9 | 0.85 | 0.95 | 16 |
| Korea, Democratic People's Republic of | SEAR | low | 0.9 | 0.85 | 0.95 | 16 |
| Paraguay | AMR | umi | 0.84 | 0.79 | 0.89 | 16 |
| Palestine, State of | EMR | lmi | 0.9 | 0.85 | 0.95 | 16 (SEAR average) * |
| Rwanda | AFR | low | 0.78 | 0.73 | 0.83 | 16 |
| Sudan | EMR | low | 0.78 | 0.73 | 0.83 | 16 (AFR average) * |
| Senegal | AFR | lmi | 0.78 | 0.73 | 0.83 | 16 |
| Solomon Islands | WPR | lmi | 0.9 | 0.85 | 0.95 | 16 |
| Sierra Leone | AFR | low | 0.78 | 0.73 | 0.83 | 16 |
| El Salvador | AMR | umi | 0.84 | 0.79 | 0.89 | 16 |
| Somalia | EMR | low | 0.78 | 0.73 | 0.83 | 16 (AFR average) * |
| Serbia | EUR | umi | 0.91 | 0.86 | 0.96 | 16 |
| South Sudan | AFR | low | 0.78 | 0.73 | 0.83 | 16 |
| Sao Tome and Principe | AFR | lmi | 0.78 | 0.73 | 0.83 | 16 |
| Swaziland | AFR | | 0.78 | 0.73 | 0.83 | 16 |
| Syrian Arab Republic | EMR | low | 0.9 | 0.85 | 0.95 | 16 (SEAR average) * |
| Chad | AFR | low | 0.78 | 0.73 | 0.83 | 16 |
| Togo | AFR | low | 0.78 | 0.73 | 0.83 | 16 |
| Thailand | SEAR | umi | 0.9 | 0.85 | 0.95 | 16 (SEAR average) * |
| Tajikistan | EUR | lmi | 0.91 | 0.86 | 0.96 | 16 |
| Turkmenistan | EUR | umi | 0.91 | 0.86 | 0.96 | 16 |
| Timor-Leste | SEAR | lmi | 0.9 | 0.85 | 0.95 | 16 |

| | | | | | | |
|---|---|---|---|---|---|---|
| Tonga | WPR | umi | 0.9 | 0.85 | 0.95 | 16 |
| Tunisia | EMR | lmi | 0.78 | 0.73 | 0.83 | 16 (AFR average) * |
| Tuvalu | WPR | umi | 0.9 | 0.85 | 0.95 | 16 |
| Tanzania, United Republic of | AFR | lmi | 0.78 | 0.73 | 0.83 | 16 |
| Uganda | AFR | low | 0.78 | 0.73 | 0.83 | 16 |
| Ukraine | EUR | lmi | 0.91 | 0.86 | 0.96 | 16 |
| Uzbekistan | EUR | lmi | 0.91 | 0.86 | 0.96 | 16 |
| St. Vincent & the Grenadines | AMR | umi | 0.84 | 0.79 | 0.89 | 16 |
| Venezuela, Bolivarian Republic of | AMR | | 0.84 | 0.79 | 0.89 | 16 |
| Viet Nam | WPR | lmi | 0.9 | 0.85 | 0.95 | 16 |
| Vanuatu | WPR | lmi | 0.9 | 0.85 | 0.95 | 16 |
| Samoa | WPR | | 0.9 | 0.85 | 0.95 | 16 |
| Kosovo | EUR | umi | 0.91 | 0.86 | 0.96 | 16 |
| Yemen | EMR | low | 0.9 | 0.85 | 0.95 | 16 (SEAR average) * |
| South Africa | AFR | umi | 0.78 | 0.73 | 0.83 | 16 |
| Zambia | AFR | lmi | 0.78 | 0.73 | 0.83 | 16 |
| Zimbabwe | AFR | lmi | 0.78 | 0.73 | 0.83 | 16 |

**Table S3:** Type distribution assumptions for anal cancer.

**Note:** Asterisks (*) denote instances where the authors performed calculations to determine the average. AFR= African Region; AMR= Region of the Americas; EMR= Eastern Mediterranean Region; EUR= European Region; lb= lower bound; lmi= lower-middle income; ub= upper bound; umi= upper-middle income; WPR= Western Pacific Region.

| Country Name | Region | Income Level | Type distribution mean | Lower Bound | Upper Bound | Source |
|---|---|---|---|---|---|---|
| Afghanistan | EMR | low | 0.95 | 0.19 | 1 | 16 (SEAR average) * |
| Angola | AFR | lmi | 0.13 | 0 | 0.25 | 17, 18 |
| Albania | EUR | umi | 0.92 | 0.16 | 1 | 16 |
| Armenia | EUR | umi | 0.92 | 0.16 | 1 | 16 |
| Azerbaijan | EUR | umi | 0.92 | 0.16 | 1 | 16 |
| Burundi | AFR | low | 0.13 | 0 | 0.25 | 17, 18 |
| Benin | AFR | lmi | 0.13 | 0 | 0.25 | 17, 18 |
| Burkina Faso | AFR | low | 0.13 | 0 | 0.25 | 17, 18 |
| Bangladesh | SEAR | lmi | 0.95 | 0.19 | 1 | 16 |
| Bosnia and Herzegovina | EUR | umi | 0.92 | 0.16 | 1 | 16 |
| Belarus | EUR | umi | 0.92 | 0.16 | 1 | 16 |
| Belize | AMR | umi | 0.81 | 0.05 | 1 | 16 |
| Bolivia, Plurinational State of | AMR | lmi | 0.81 | 0.05 | 1 | 16 |
| Bhutan | SEAR | lmi | 0.95 | 0.19 | 1 | 16 |
| Central African Republic | AFR | low | 0.13 | 0 | 0.25 | 17, 18 |
| China (Asia) | WPR | umi | 0.95 | 0.19 | 1 | 16 |
| Cote d'Ivoire | AFR | lmi | 0.13 | 0 | 0.25 | 17, 18 |
| Cameroon | AFR | lmi | 0.13 | 0 | 0.25 | 17, 18 |
| Congo, the Democratic Republic of the | AFR | low | 0.13 | 0 | 0.25 | 17, 18 |
| Congo | AFR | lmi | 0.13 | 0 | 0.25 | 17, 18 |
| Colombia | AMR | umi | 0.81 | 0.05 | 1 | 16 |
| Comoros | AFR | lmi | 0.13 | 0 | 0.25 | 17, 18 |
| Cabo Verde | AFR | lmi | 0.13 | 0 | 0.25 | 17, 18 |
| Cuba | AMR | umi | 0.81 | 0.05 | 1 | 16 |
| Djibouti | EMR | lmi | 0.13 | 0 | 0.25 | 17, 18 (AFR average) * |
| Dominica | AMR | umi | 0.81 | 0.05 | 1 | 16 |
| Algeria | AFR | lmi | 0.13 | 0 | 0.25 | 17, 18 |
| Ecuador | AMR | umi | 0.81 | 0.05 | 1 | 16 |
| Egypt | EMR | lmi | 0.13 | 0 | 0.25 | 17, 18 (AFR average) * |
| Eritrea | AFR | low | 0.13 | 0 | 0.25 | 17, 18 |
| Ethiopia | AFR | low | 0.13 | 0 | 0.25 | 17, 18 |
| Fiji | WPR | umi | 0.95 | 0.19 | 1 | 16 |
| Micronesia, Federated States of | WPR | lmi | 0.95 | 0.19 | 1 | 16 |

| | | | | | | |
|---|---|---|---|---|---|---|
| Georgia | EUR | umi | 0.92 | 0.16 | 1 | 16 |
| Ghana | AFR | lmi | 0.13 | 0 | 0.25 | 17, 18 |
| Guinea | AFR | lmi | 0.13 | 0 | 0.25 | 17, 18 |
| Gambia | AFR | low | 0.13 | 0 | 0.25 | 17, 18 |
| Guinea-Bissau | AFR | low | 0.13 | 0 | 0.25 | 17, 18 |
| Grenada | AMR | umi | 0.81 | 0.05 | 1 | 16 |
| Guatemala | AMR | umi | 0.81 | 0.05 | 1 | 16 |
| Guyana | AMR | | 0.81 | 0.05 | 1 | 16 |
| Honduras | AMR | lmi | 0.81 | 0.05 | 1 | 16 |
| Haiti | AMR | lmi | 0.81 | 0.05 | 1 | 16 |
| Indonesia | SEAR | umi | 0.95 | 0.19 | 1 | 16 |
| India | SEAR | lmi | 0.95 | 0.19 | 1 | 16 |
| Iran, Islamic Republic of | EMR | lmi | 0.95 | 0.19 | 1 | 16 (SEAR average) * |
| Iraq | EMR | umi | 0.95 | 0.19 | 1 | 16 (SEAR average) * |
| Jamaica | AMR | umi | 0.81 | 0.05 | 1 | 16 |
| Jordan | EMR | lmi | 0.95 | 0.19 | 1 | 16 |
| Kenya | AFR | lmi | 0.13 | 0 | 0.25 | 17, 18 |
| Kyrgyzstan | EUR | | 0.92 | 0.16 | 1 | 16 |
| Cambodia | WPR | lmi | 0.95 | 0.19 | 1 | 16 |
| Kiribati | WPR | lmi | 0.95 | 0.19 | 1 | 16 |
| Lao People's Democratic Republic | WPR | lmi | 0.95 | 0.19 | 1 | 16 |
| Liberia | AFR | low | 0.13 | 0 | 0.25 | 17, 18 |
| St. Lucia | AMR | umi | 0.81 | 0.05 | 1 | 16 |
| Sri Lanka | SEAR | lmi | 0.95 | 0.19 | 1 | 16 |
| Lesotho | AFR | lmi | 0.13 | 0 | 0.25 | 17, 18 |
| Morocco | EMR | lmi | 0.13 | 0 | 0.25 | 17, 18 (AFR average) * |
| Moldova, Republic of | EUR | umi | 0.92 | 0.16 | 1 | 16 |
| Madagascar | AFR | low | 0.13 | 0 | 0.25 | 17, 18 |
| Maldives | SEAR | umi | 0.95 | 0.19 | 1 | 16 |
| Marshall Islands | WPR | umi | 0.95 | 0.19 | 1 | 16 |
| Macedonia, the former Yugoslav Republic of | EUR | | 0.92 | 0.16 | 1 | 16 |
| Mali | AFR | low | 0.13 | 0 | 0.25 | 17, 18 |
| Myanmar | SEAR | lmi | 0.95 | 0.19 | 1 | 16 |
| Mongolia | WPR | lmi | 0.95 | 0.19 | 1 | 16 |
| Mozambique | AFR | low | 0.13 | 0 | 0.25 | 17, 18 |
| Mauritania | AFR | lmi | 0.13 | 0 | 0.25 | 17, 18 |
| Malawi | AFR | low | 0.13 | 0 | 0.25 | 17, 18 |

| | | | | | | |
|---|---|---|---|---|---|---|
| Namibia | AFR | umi | 0.13 | 0 | 0.25 | 17, 18 |
| Niger | AFR | low | 0.13 | 0 | 0.25 | 17, 18 |
| Nigeria | AFR | lmi | 0.13 | 0 | 0.25 | 17, 18 |
| Nicaragua | AMR | lmi | 0.81 | 0.05 | 1 | 16 |
| Nepal | SEAR | lmi | 0.95 | 0.19 | 1 | 16 |
| Pakistan | EMR | lmi | 0.95 | 0.19 | 1 | 16 (SEAR average) * |
| Peru | AMR | umi | 0.81 | 0.05 | 1 | 16 |
| Philippines (Asia) | WPR | lmi | 0.95 | 0.19 | 1 | 16 |
| Papua New Guinea | WPR | lmi | 0.95 | 0.19 | 1 | 16 |
| Korea, Democratic People's Republic of | SEAR | low | 0.95 | 0.19 | 1 | 16 |
| Paraguay | AMR | umi | 0.81 | 0.05 | 1 | 16 |
| Palestine, State of | EMR | lmi | 0.95 | 0.19 | 1 | 16 (SEAR average) * |
| Rwanda | AFR | low | 0.13 | 0 | 0.25 | 17, 18 |
| Sudan | EMR | low | 0.13 | 0 | 0.25 | 17,18 (AFR average) * |
| Senegal | AFR | lmi | 0.13 | 0 | 0.25 | 17, 18 |
| Solomon Islands | WPR | lmi | 0.95 | 0.19 | 1 | 16 |
| Sierra Leone | AFR | low | 0.13 | 0 | 0.25 | 17, 18 |
| El Salvador | AMR | umi | 0.81 | 0.05 | 1 | 16 |
| Somalia | EMR | low | 0.13 | 0 | 0.25 | 17, 18 (AFR average) * |
| Serbia | EUR | umi | 0.92 | 0.16 | 1 | 16 |
| South Sudan | AFR | low | 0.13 | 0 | 0.25 | 17, 18 |
| Sao Tome and Principe | AFR | lmi | 0.13 | 0 | 0.25 | 17, 18 |
| Swaziland | AFR | | 0.13 | 0 | 0.25 | 17, 18 |
| Syrian Arab Republic | EMR | low | 0.95 | 0.19 | 1 | 16 (SEAR average) * |
| Chad | AFR | low | 0.13 | 0 | 0.25 | 17, 18 |
| Togo | AFR | low | 0.13 | 0 | 0.25 | 17, 18 |
| Thailand | SEAR | umi | 0.95 | 0.19 | 1 | 16 (SEAR average) * |
| Tajikistan | EUR | lmi | 0.92 | 0.16 | 1 | 16 |
| Turkmenistan | EUR | umi | 0.92 | 0.16 | 1 | 16 |
| Timor-Leste | SEAR | lmi | 0.95 | 0.19 | 1 | 16 |
| Tonga | WPR | umi | 0.95 | 0.19 | 1 | 16 |
| Tunisia | EMR | lmi | 0.13 | 0 | 0.25 | 17, 18 (AFR average) * |
| Tuvalu | WPR | umi | 0.95 | 0.19 | 1 | 16 |
| Tanzania, United Republic of | AFR | lmi | 0.13 | 0 | 0.25 | 17, 18 |
| Uganda | AFR | low | 0.13 | 0 | 0.25 | 17, 18 |

| | | | | | | |
|---|---|---|---|---|---|---|
| Ukraine | EUR | lmi | 0.92 | 0.16 | 1 | 16 |
| Uzbekistan | EUR | lmi | 0.92 | 0.16 | 1 | 16 |
| St. Vincent & the Grenadines | AMR | umi | 0.81 | 0.05 | 1 | 16 |
| Venezuela, Bolivarian Republic of | AMR | | 0.81 | 0.05 | 1 | 16 |
| Viet Nam | WPR | lmi | 0.95 | 0.19 | 1 | 16 |
| Vanuatu | WPR | lmi | 0.95 | 0.19 | 1 | 16 |
| Samoa | WPR | | 0.95 | 0.19 | 1 | 16 |
| Kosovo | EUR | umi | 0.92 | 0.16 | 1 | 16 |
| Yemen | EMR | low | 0.95 | 0.19 | 1 | 16 (SEAR average) * |
| South Africa | AFR | umi | 0.13 | 0 | 0.25 | 17, 18 |
| Zambia | AFR | lmi | 0.13 | 0 | 0.25 | 17, 18 |
| Zimbabwe | AFR | lmi | 0.13 | 0 | 0.25 | 17, 18 |

**Table S4:** Type distribution assumptions for oropharyngeal cancer.

**Note:** Asterisks (*) denote instances where the authors performed calculations to determine the average. For sources 17 and 18 the lower bound was assumed to be zero per de Sanjosé, et al. (2019), and the upper bound was set to 25% per Castellsagué, et al. (2016), reflecting HPV-DNA positivity in oropharyngeal cancers globally. AFR= African Region; AMR= Region of the Americas; EMR= Eastern Mediterranean Region; EUR= European Region; lb= lower bound; lmi= lower-middle income; ub= upper bound; umi= upper-middle income; WPR= Western Pacific Region.

| Country Name | Region | Income Level | Type distribution mean | Lower Bound | Upper Bound | Source |
|---|---|---|---|---|---|---|
| Afghanistan | EMR | low | 0.6 | 0.48 | 0.72 | 16 (SEAR average) * |
| Angola | AFR | lmi | 0.53 | 0.41 | 0.65 | 16 |
| Albania | EUR | umi | 0.72 | 0.60 | 0.84 | 16 |
| Armenia | EUR | umi | 0.72 | 0.60 | 0.84 | 16 |
| Azerbaijan | EUR | umi | 0.72 | 0.60 | 0.84 | 16 |
| Burundi | AFR | low | 0.53 | 0.41 | 0.65 | 16 |
| Benin | AFR | lmi | 0.53 | 0.41 | 0.65 | 16 |
| Burkina Faso | AFR | low | 0.53 | 0.41 | 0.65 | 16 |
| Bangladesh | SEAR | lmi | 0.6 | 0.48 | 0.72 | 16 |
| Bosnia and Herzegovina | EUR | umi | 0.72 | 0.60 | 0.84 | 16 |
| Belarus | EUR | umi | 0.72 | 0.60 | 0.84 | 16 |
| Belize | AMR | umi | 0.6 | 0.48 | 0.72 | 16 |
| Bolivia, Plurinational State of | AMR | lmi | 0.6 | 0.48 | 0.72 | 16 |
| Bhutan | SEAR | lmi | 0.6 | 0.48 | 0.72 | 16 |
| Central African Republic | AFR | low | 0.53 | 0.41 | 0.65 | 16 |
| China (Asia) | WPR | umi | 0.6 | 0.48 | 0.72 | 16 |
| Cote d'Ivoire | AFR | lmi | 0.53 | 0.41 | 0.65 | 16 |
| Cameroon | AFR | lmi | 0.53 | 0.41 | 0.65 | 16 |
| Congo, the Democratic Republic of the | AFR | low | 0.53 | 0.41 | 0.65 | 16 |
| Congo | AFR | lmi | 0.53 | 0.41 | 0.65 | 16 |
| Colombia | AMR | umi | 0.6 | 0.48 | 0.72 | 16 |
| Comoros | AFR | lmi | 0.53 | 0.41 | 0.65 | 16 |
| Cabo Verde | AFR | lmi | 0.53 | 0.41 | 0.65 | 16 |
| Cuba | AMR | umi | 0.6 | 0.48 | 0.72 | 16 |
| Djibouti | EMR | lmi | 0.53 | 0.41 | 0.65 | 16 (AFR average) * |
| Dominica | AMR | umi | 0.6 | 0.48 | 0.72 | 16 |
| Algeria | AFR | lmi | 0.53 | 0.41 | 0.65 | 16 |
| Ecuador | AMR | umi | 0.6 | 0.48 | 0.72 | 16 |
| Egypt | EMR | lmi | 0.53 | 0.41 | 0.65 | 16 (AFR average) * |
| Eritrea | AFR | low | 0.53 | 0.41 | 0.65 | 16 |
| Ethiopia | AFR | low | 0.53 | 0.41 | 0.65 | 16 |
| Fiji | WPR | umi | 0.6 | 0.48 | 0.72 | 16 |

| | | | | | | |
|---|---|---|---|---|---|---|
| Micronesia, Federated States of | WPR | lmi | 0.6 | 0.48 | 0.72 | 16 |
| Georgia | EUR | umi | 0.72 | 0.60 | 0.84 | 16 |
| Ghana | AFR | lmi | 0.53 | 0.41 | 0.65 | 16 |
| Guinea | AFR | lmi | 0.53 | 0.41 | 0.65 | 16 |
| Gambia | AFR | low | 0.53 | 0.41 | 0.65 | 16 |
| Guinea-Bissau | AFR | low | 0.53 | 0.41 | 0.65 | 16 |
| Grenada | AMR | umi | 0.6 | 0.48 | 0.72 | 16 |
| Guatemala | AMR | umi | 0.6 | 0.48 | 0.72 | 16 |
| Guyana | AMR | | 0.6 | 0.48 | 0.72 | 16 |
| Honduras | AMR | lmi | 0.6 | 0.48 | 0.72 | 16 |
| Haiti | AMR | lmi | 0.6 | 0.48 | 0.72 | 16 |
| Indonesia | SEAR | umi | 0.6 | 0.48 | 0.72 | 16 |
| India | SEAR | lmi | 0.6 | 0.48 | 0.72 | 16 |
| Iran, Islamic Republic of | EMR | lmi | 0.6 | 0.48 | 0.72 | 16 (SEAR average) * |
| Iraq | EMR | umi | 0.6 | 0.48 | 0.72 | 16 (SEAR average) * |
| Jamaica | AMR | umi | 0.6 | 0.48 | 0.72 | 16 |
| Jordan | EMR | lmi | 0.6 | 0.48 | 0.72 | 16 |
| Kenya | AFR | lmi | 0.53 | 0.41 | 0.65 | 16 |
| Kyrgyzstan | EUR | | 0.72 | 0.60 | 0.84 | 16 |
| Cambodia | WPR | lmi | 0.6 | 0.48 | 0.72 | 16 |
| Kiribati | WPR | lmi | 0.6 | 0.48 | 0.72 | 16 |
| Lao People's Democratic Republic | WPR | lmi | 0.6 | 0.48 | 0.72 | 16 |
| Liberia | AFR | low | 0.53 | 0.41 | 0.65 | 16 |
| St. Lucia | AMR | umi | 0.6 | 0.48 | 0.72 | 16 |
| Sri Lanka | SEAR | lmi | 0.6 | 0.48 | 0.72 | 16 |
| Lesotho | AFR | lmi | 0.53 | 0.41 | 0.65 | 16 |
| Morocco | EMR | lmi | 0.53 | 0.41 | 0.65 | 16 (AFR average) * |
| Moldova, Republic of | EUR | umi | 0.72 | 0.60 | 0.84 | 16 |
| Madagascar | AFR | low | 0.53 | 0.41 | 0.65 | 16 |
| Maldives | SEAR | umi | 0.6 | 0.48 | 0.72 | 16 |
| Marshall Islands | WPR | umi | 0.6 | 0.48 | 0.72 | 16 |
| Macedonia, the former Yugoslav Republic of | EUR | | 0.72 | 0.60 | 0.84 | 16 |
| Mali | AFR | low | 0.53 | 0.41 | 0.65 | 16 |

| | | | | | | |
|---|---|---|---|---|---|---|
| Myanmar | SEAR | lmi | 0.6 | 0.48 | 0.72 | 16 |
| Mongolia | WPR | lmi | 0.6 | 0.48 | 0.72 | 16 |
| Mozambique | AFR | low | 0.53 | 0.41 | 0.65 | 16 |
| Mauritania | AFR | lmi | 0.53 | 0.41 | 0.65 | 16 |
| Malawi | AFR | low | 0.53 | 0.41 | 0.65 | 16 |
| Namibia | AFR | umi | 0.53 | 0.41 | 0.65 | 16 |
| Niger | AFR | low | 0.53 | 0.41 | 0.65 | 16 |
| Nigeria | AFR | lmi | 0.53 | 0.41 | 0.65 | 16 |
| Nicaragua | AMR | lmi | 0.6 | 0.48 | 0.72 | 16 |
| Nepal | SEAR | lmi | 0.6 | 0.48 | 0.72 | 16 |
| Pakistan | EMR | lmi | 0.6 | 0.48 | 0.72 | 16 (SEAR average) * |
| Peru | AMR | umi | 0.6 | 0.48 | 0.72 | 16 |
| Philippines (Asia) | WPR | lmi | 0.6 | 0.48 | 0.72 | 16 |
| Papua New Guinea | WPR | lmi | 0.6 | 0.48 | 0.72 | 16 |
| Korea, Democratic People's Republic of | SEAR | low | 0.6 | 0.48 | 0.72 | 16 |
| Paraguay | AMR | umi | 0.6 | 0.48 | 0.72 | 16 |
| Palestine, State of | EMR | lmi | 0.6 | 0.48 | 0.72 | 16 (SEAR average) * |
| Rwanda | AFR | low | 0.53 | 0.41 | 0.65 | 16 |
| Sudan | EMR | low | 0.53 | 0.41 | 0.65 | 16 (AFR average) * |
| Senegal | AFR | lmi | 0.53 | 0.41 | 0.65 | 16 |
| Solomon Islands | WPR | lmi | 0.6 | 0.48 | 0.72 | 16 |
| Sierra Leone | AFR | low | 0.53 | 0.41 | 0.65 | 16 |
| El Salvador | AMR | umi | 0.6 | 0.48 | 0.72 | 16 |
| Somalia | EMR | low | 0.53 | 0.41 | 0.65 | 16 (AFR average) * |
| Serbia | EUR | umi | 0.72 | 0.60 | 0.84 | 16 |
| South Sudan | AFR | low | 0.53 | 0.41 | 0.65 | 16 |
| Sao Tome and Principe | AFR | lmi | 0.53 | 0.41 | 0.65 | 16 |
| Swaziland | AFR | | 0.53 | 0.41 | 0.65 | 16 |
| Syrian Arab Republic | EMR | low | 0.6 | 0.48 | 0.72 | 16 (SEAR average) * |
| Chad | AFR | low | 0.53 | 0.41 | 0.65 | 16 |
| Togo | AFR | low | 0.53 | 0.41 | 0.65 | 16 |
| Thailand | SEAR | umi | 0.6 | 0.48 | 0.72 | 16 (SEAR average) * |
| Tajikistan | EUR | lmi | 0.72 | 0.60 | 0.84 | 16 |
| Turkmenistan | EUR | umi | 0.72 | 0.60 | 0.84 | 16 |
| Timor-Leste | SEAR | lmi | 0.6 | 0.48 | 0.72 | 16 |

| | | | | | | |
|---|---|---|---|---|---|---|
| Tonga | WPR | umi | 0.6 | 0.48 | 0.72 | 16 |
| Tunisia | EMR | lmi | 0.53 | 0.41 | 0.65 | 16 (AFR average) * |
| Tuvalu | WPR | umi | 0.6 | 0.48 | 0.72 | 16 |
| Tanzania, United Republic of | AFR | lmi | 0.53 | 0.41 | 0.65 | 16 |
| Uganda | AFR | low | 0.53 | 0.41 | 0.65 | 16 |
| Ukraine | EUR | lmi | 0.72 | 0.60 | 0.84 | 16 |
| Uzbekistan | EUR | lmi | 0.72 | 0.60 | 0.84 | 16 |
| St. Vincent & the Grenadines | AMR | umi | 0.6 | 0.48 | 0.72 | 16 |
| Venezuela, Bolivarian Republic of | AMR | | 0.6 | 0.48 | 0.72 | 16 |
| Viet Nam | WPR | lmi | 0.6 | 0.48 | 0.72 | 16 |
| Vanuatu | WPR | lmi | 0.6 | 0.48 | 0.72 | 16 |
| Samoa | WPR | | 0.6 | 0.48 | 0.72 | 16 |
| Kosovo | EUR | umi | 0.72 | 0.60 | 0.84 | 16 |
| Yemen | EMR | low | 0.6 | 0.48 | 0.72 | 16 (SEAR average) * |
| South Africa | AFR | umi | 0.53 | 0.41 | 0.65 | 16 |
| Zambia | AFR | lmi | 0.53 | 0.41 | 0.65 | 16 |
| Zimbabwe | AFR | lmi | 0.53 | 0.41 | 0.65 | 16 |

**Table S5:** Type distribution assumptions for vaginal cancer.

**Note:** Asterisks (*) denote instances where the authors performed calculations to determine the average. AFR= African Region; AMR= Region of the Americas; EMR= Eastern Mediterranean Region; EUR= European Region; lb= lower bound; lmi= lower-middle income; ub= upper bound; umi= upper-middle income; WPR= Western Pacific Region.

| Country Name | Region | Income Level | Type distribution mean | Lower Bound | Upper Bound | Source |
|---|---|---|---|---|---|---|
| Afghanistan | EMR | low | 0.76 | 0.61 | 0.91 | 16 (SEAR average) * |
| Angola | AFR | lmi | 0.92 | 0.77 | 1.00 | 16 |
| Albania | EUR | umi | 0.82 | 0.67 | 0.97 | 16 |
| Armenia | EUR | umi | 0.82 | 0.67 | 0.97 | 16 |
| Azerbaijan | EUR | umi | 0.82 | 0.67 | 0.97 | 16 |
| Burundi | AFR | low | 0.92 | 0.77 | 1.00 | 16 |
| Benin | AFR | lmi | 0.92 | 0.77 | 1.00 | 16 |
| Burkina Faso | AFR | low | 0.92 | 0.77 | 1.00 | 16 |
| Bangladesh | SEAR | lmi | 0.76 | 0.61 | 0.91 | 16 |
| Bosnia and Herzegovina | EUR | umi | 0.82 | 0.67 | 0.97 | 16 |
| Belarus | EUR | umi | 0.82 | 0.67 | 0.97 | 16 |
| Belize | AMR | umi | 0.71 | 0.56 | 0.86 | 16 |
| Bolivia, Plurinational State of | AMR | lmi | 0.71 | 0.56 | 0.86 | 16 |
| Bhutan | SEAR | lmi | 0.76 | 0.61 | 0.91 | 16 |
| Central African Republic | AFR | low | 0.92 | 0.77 | 1.00 | 16 |
| China (Asia) | WPR | umi | 0.76 | 0.61 | 0.91 | 16 |
| Cote d'Ivoire | AFR | lmi | 0.92 | 0.77 | 1.00 | 16 |
| Cameroon | AFR | lmi | 0.92 | 0.77 | 1.00 | 16 |
| Congo, the Democratic Republic of the | AFR | low | 0.92 | 0.77 | 1.00 | 16 |
| Congo | AFR | lmi | 0.92 | 0.77 | 1.00 | 16 |
| Colombia | AMR | umi | 0.71 | 0.56 | 0.86 | 16 |
| Comoros | AFR | lmi | 0.92 | 0.77 | 1.00 | 16 |
| Cabo Verde | AFR | lmi | 0.92 | 0.77 | 1.00 | 16 |
| Cuba | AMR | umi | 0.71 | 0.56 | 0.86 | 16 |
| Djibouti | EMR | lmi | 0.92 | 0.77 | 1.00 | 16 (AFR average) * |
| Dominica | AMR | umi | 0.71 | 0.56 | 0.86 | 16 |
| Algeria | AFR | lmi | 0.92 | 0.77 | 1.00 | 16 |
| Ecuador | AMR | umi | 0.71 | 0.56 | 0.86 | 16 |
| Egypt | EMR | lmi | 0.92 | 0.77 | 1.00 | 16 (AFR average) * |
| Eritrea | AFR | low | 0.92 | 0.77 | 1.00 | 16 |
| Ethiopia | AFR | low | 0.92 | 0.77 | 1.00 | 16 |
| Fiji | WPR | umi | 0.76 | 0.61 | 0.91 | 16 |

| | | | | | | |
|---|---|---|---|---|---|---|
| Micronesia, Federated States of | WPR | lmi | 0.76 | 0.61 | 0.91 | 16 |
| Georgia | EUR | umi | 0.82 | 0.67 | 0.97 | 16 |
| Ghana | AFR | lmi | 0.92 | 0.77 | 1.00 | 16 |
| Guinea | AFR | lmi | 0.92 | 0.77 | 1.00 | 16 |
| Gambia | AFR | low | 0.92 | 0.77 | 1.00 | 16 |
| Guinea-Bissau | AFR | low | 0.92 | 0.77 | 1.00 | 16 |
| Grenada | AMR | umi | 0.71 | 0.56 | 0.86 | 16 |
| Guatemala | AMR | umi | 0.71 | 0.56 | 0.86 | 16 |
| Guyana | AMR | | 0.71 | 0.56 | 0.86 | 16 |
| Honduras | AMR | lmi | 0.71 | 0.56 | 0.86 | 16 |
| Haiti | AMR | lmi | 0.71 | 0.56 | 0.86 | 16 |
| Indonesia | SEAR | umi | 0.76 | 0.61 | 0.91 | 16 |
| India | SEAR | lmi | 0.76 | 0.61 | 0.91 | 16 |
| Iran, Islamic Republic of | EMR | lmi | 0.92 | 0.77 | 1.00 | 16 (SEAR average) * |
| Iraq | EMR | umi | 0.76 | 0.61 | 0.91 | 16 (SEAR average) * |
| Jamaica | AMR | umi | 0.71 | 0.56 | 0.86 | 16 |
| Jordan | EMR | lmi | 0.76 | 0.61 | 0.91 | 16 |
| Kenya | AFR | lmi | 0.92 | 0.77 | 1.00 | 16 |
| Kyrgyzstan | EUR | | 0.82 | 0.67 | 0.97 | 16 |
| Cambodia | WPR | lmi | 0.76 | 0.61 | 0.91 | 16 |
| Kiribati | WPR | lmi | 0.76 | 0.61 | 0.91 | 16 |
| Lao People's Democratic Republic | WPR | lmi | 0.76 | 0.61 | 0.91 | 16 |
| Liberia | AFR | low | 0.92 | 0.77 | 1.00 | 16 |
| St. Lucia | AMR | umi | 0.71 | 0.56 | 0.86 | 16 |
| Sri Lanka | SEAR | lmi | 0.76 | 0.61 | 0.91 | 16 |
| Lesotho | AFR | lmi | 0.92 | 0.77 | 1.00 | 16 |
| Morocco | EMR | lmi | 0.92 | 0.77 | 1.00 | 16 (AFR average) * |
| Moldova, Republic of | EUR | umi | 0.82 | 0.67 | 0.97 | 16 |
| Madagascar | AFR | low | 0.92 | 0.77 | 1.00 | 16 |
| Maldives | SEAR | umi | 0.76 | 0.61 | 0.91 | 16 |
| Marshall Islands | WPR | umi | 0.76 | 0.61 | 0.91 | 16 |
| Macedonia, the former Yugoslav Republic of | EUR | | 0.82 | 0.67 | 0.97 | 16 |
| Mali | AFR | low | 0.92 | 0.77 | 1.00 | 16 |

| | | | | | | |
|---|---|---|---|---|---|---|
| Myanmar | SEAR | lmi | 0.76 | 0.61 | 0.91 | 16 |
| Mongolia | WPR | lmi | 0.76 | 0.61 | 0.91 | 16 |
| Mozambique | AFR | low | 0.92 | 0.77 | 1.00 | 16 |
| Mauritania | AFR | lmi | 0.92 | 0.77 | 1.00 | 16 |
| Malawi | AFR | low | 0.92 | 0.77 | 1.00 | 16 |
| Namibia | AFR | umi | 0.92 | 0.77 | 1.00 | 16 |
| Niger | AFR | low | 0.92 | 0.77 | 1.00 | 16 |
| Nigeria | AFR | lmi | 0.92 | 0.77 | 1.00 | 16 |
| Nicaragua | AMR | lmi | 0.71 | 0.56 | 0.86 | 16 |
| Nepal | SEAR | lmi | 0.76 | 0.61 | 0.91 | 16 |
| Pakistan | EMR | lmi | 0.76 | 0.61 | 0.91 | 16 (SEAR average) * |
| Peru | AMR | umi | 0.71 | 0.56 | 0.86 | 16 |
| Philippines (Asia) | WPR | lmi | 0.76 | 0.61 | 0.91 | 16 |
| Papua New Guinea | WPR | lmi | 0.76 | 0.61 | 0.91 | 16 |
| Korea, Democratic People's Republic of | SEAR | low | 0.76 | 0.61 | 0.91 | 16 |
| Paraguay | AMR | umi | 0.71 | 0.56 | 0.86 | 16 |
| Palestine, State of | EMR | lmi | 0.76 | 0.61 | 0.91 | 16 (SEAR average) * |
| Rwanda | AFR | low | 0.92 | 0.77 | 1.00 | 16 |
| Sudan | EMR | low | 0.92 | 0.77 | 1.00 | 16 (AFR average) * |
| Senegal | AFR | lmi | 0.92 | 0.77 | 1.00 | 16 |
| Solomon Islands | WPR | lmi | 0.76 | 0.61 | 0.91 | 16 |
| Sierra Leone | AFR | low | 0.92 | 0.77 | 1.00 | 16 |
| El Salvador | AMR | umi | 0.71 | 0.56 | 0.86 | 16 |
| Somalia | EMR | low | 0.92 | 0.77 | 1.00 | 16 (AFR average) * |
| Serbia | EUR | umi | 0.82 | 0.67 | 0.97 | 16 |
| South Sudan | AFR | low | 0.92 | 0.77 | 1.00 | 16 |
| Sao Tome and Principe | AFR | lmi | 0.92 | 0.77 | 1.00 | 16 |
| Swaziland | AFR | | 0.92 | 0.77 | 1.00 | 16 |
| Syrian Arab Republic | EMR | low | 0.76 | 0.61 | 0.91 | 16 (SEAR average) * |
| Chad | AFR | low | 0.92 | 0.77 | 1.00 | 16 |
| Togo | AFR | low | 0.92 | 0.77 | 1.00 | 16 |
| Thailand | SEAR | umi | 0.76 | 0.61 | 0.91 | 16 (SEAR average) * |
| Tajikistan | EUR | lmi | 0.82 | 0.67 | 0.97 | 16 |
| Turkmenistan | EUR | umi | 0.82 | 0.67 | 0.97 | 16 |
| Timor-Leste | SEAR | lmi | 0.76 | 0.61 | 0.91 | 16 |

| | | | | | | |
|---|---|---|---|---|---|---|
| Tonga | WPR | umi | 0.76 | 0.61 | 0.91 | 16 |
| Tunisia | EMR | lmi | 0.92 | 0.77 | 1.00 | 16 (AFR average) * |
| Tuvalu | WPR | umi | 0.76 | 0.61 | 0.91 | 16 |
| Tanzania, United Republic of | AFR | lmi | 0.92 | 0.77 | 1.00 | 16 |
| Uganda | AFR | low | 0.92 | 0.77 | 1.00 | 16 |
| Ukraine | EUR | lmi | 0.82 | 0.67 | 0.97 | 16 |
| Uzbekistan | EUR | lmi | 0.82 | 0.67 | 0.97 | 16 |
| St. Vincent & the Grenadines | AMR | umi | 0.71 | 0.56 | 0.86 | 16 |
| Venezuela, Bolivarian Republic of | AMR | | 0.71 | 0.56 | 0.86 | 16 |
| Viet Nam | WPR | lmi | 0.76 | 0.61 | 0.91 | 16 |
| Vanuatu | WPR | lmi | 0.76 | 0.61 | 0.91 | 16 |
| Samoa | WPR | | 0.76 | 0.61 | 0.91 | 16 |
| Kosovo | EUR | umi | 0.82 | 0.67 | 0.97 | 16 |
| Yemen | EMR | low | 0.76 | 0.61 | 0.91 | 16 (SEAR average) * |
| South Africa | AFR | umi | 0.92 | 0.77 | 1.00 | 16 |
| Zambia | AFR | lmi | 0.92 | 0.77 | 1.00 | 16 |
| Zimbabwe | AFR | lmi | 0.92 | 0.77 | 1.00 | 16 |

**Table S6:** Type distribution assumptions for vulvar cancer.

**Note:** Asterisks (*) denote instances where the authors performed calculations to determine the average. AFR= African Region; AMR= Region of the Americas; EMR= Eastern Mediterranean Region; EUR= European Region; lb= lower bound; lmi= lower-middle income; ub= upper bound; umi= upper-middle income; WPR= Western Pacific Region.

**CUMULATIVE CASES AVERTED BY COUNTRY ACROSS 2030–2100**

| COUNTRY | ANAL | OROPHARYNGEAL | VAGINAL | VULVAR |
|---|---|---|---|---|
| AFGHANISTAN | 6,860 (5,450–8,380) | 6,560 (4,010–8,600) | 1,970 (1,430–2,600) | 5,180 (3,850–6,610) |
| ANGOLA | 2,730 (1,880–3,770) | 9,580 (5,150–14,200) | 8,140 (5,410–11,500) | 11,100 (7,290–15,500) |
| ALBANIA | 42 (25–63) | 7 (2–13) | 77 (53–105) | 225 (167–283) |
| ARMENIA | 88 (54–127) | NA | 92 (57–134) | 243 (145–354) |
| AZERBAIJAN | 928 (570–1,320) | 48 (11–87) | 33 (16–51) | 549 (353–754) |
| BURUNDI | 3,930 (2,830–4,900) | 135 (36–262) | 1,480 (1,050–1,930) | 8,570 (6,010–10,900) |
| BENIN | 1,910 (1,490–2,350) | 2,130 (1,090–2,940) | 1,630 (1,210–2,040) | 3,830 (3,010–4,830) |
| BURKINA FASO | 13,400 (9,010–18,000) | NA | 2,890 (782–5,090) | 4,670 (2,700–6,720) |
| BANGLADESH | 4,940 (3,870–6,100) | 4,240 (1,350–7,700) | 6,620 (4,900–8,560) | 13,700 (10,700–17,100) |
| BOSNIA AND HERZEGOVINA | 57 (35–81) | 101 (47–152) | 95 (66–123) | 248 (164–335) |
| BELARUS | 931 (674–1,170) | 69 (22–130) | 183 (132–240) | 1,690 (1,240–2,140) |
| BELIZE | NA | NA | NA | NA |
| BOLIVIA, PLURINATIONAL STATE OF | 401 (244–577) | 1,810 (572–3,570) | 1,300 (692–1,900) | 1,470 (840–2,090) |
| BHUTAN | NA | 313 (178–490) | NA | NA |
| CENTRAL AFRICAN REPUBLIC | 1,420 (1,040–1,810) | NA | 393 (105–692) | 2,610 (1,890–3,370) |
| CHINA | 21,600 (17,100–26,800) | 11,900 (5,570–17,800) | 18,600 (14,200–23,200) | 31,400 (24,100–39,500) |
| COTE D'IVOIRE | 7,280 (5,270–9,240) | 2,130 (1,050–3,330) | 5,970 (3,990–8,040) | 4,090 (2,790–5,390) |
| CAMEROON | 5,570 (4,170–7,030) | 483 (149–851) | 3,400 (2,290–4,460) | 6,410 (4,840–7,930) |
| CONGO, THE DEMOCRATIC REPUBLIC OF THE | 42,300 (32,700–52,300) | 443 (126–799) | 12,800 (8,980–16,500) | 63,900 (48,800–77,100) |
| CONGO | 1,750 (1,250–2,240) | 197 (62–349) | 420 (291–581) | 2,220 (1,650–2,950) |
| COLOMBIA | 10,900 (10,300–11,600) | 707 (202–1,190) | 3,520 (2,920–4,130) | 11,900 (10,800–13,000) |
| COMOROS | NA | NA | NA | 507 (396–619) |
| CABO VERDE (CAPE VERDE) | 43 (11–73) | NA | NA | 35 (10–61) |
| CUBA | 1,450 (1,020–1,850) | 89 (29–162) | 412 (288–534) | 1,280 (987–1,550) |
| DJIBOUTI | NA | NA | 51 (25–78) | 190 (125–258) |
| DOMINICA | 3 (0–5) | 1 (0–3) | 8 (4–13) | 2 (1–3) |
| ALGERIA | 2,890 (2,190–3,590) | 690 (403–960) | 1,630 (1,250–2,030) | 3,470 (2,540–4,570) |
| ECUADOR | 4,690 (3,310–6,200) | 256 (88–447) | 1,000 (708–1,320) | 4,310 (3,340–5,580) |
| EGYPT | 6,500 (5,010–7,850) | 859 (331–1,520) | 3,840 (2,710–5,110) | 15,600 (11,800–20,400) |
| ERITREA | 325 (196–464) | NA | 400 (239–576) | 1,310 (939–1,710) |

**CUMULATIVE CASES AVERTED BY COUNTRY ACROSS 2030–2100**

| COUNTRY | ANAL | OROPHARYNGEAL | VAGINAL | VULVAR |
|---|---|---|---|---|
| ETHIOPIA | 27,700 (19,600–35,000) | 4,650 (2,730–6,780) | 25,900 (18,500–33,500) | 61,300 (48,700–75,700) |
| FIJI | 247 (147–343) | 8 (1–16) | 66 (30–98) | NA |
| MICRONESIA, FEDERATED STATES OF | NA | NA | NA | NA |
| GEORGIA | 71 (42–98) | 11 (4–22) | 113 (77–155) | 773 (592–971) |
| GHANA | 10,700 (7,040–14,500) | 289 (70–652) | 909 (478–1,450) | 6,480 (4,970–8,340) |
| GUINEA | 1,250 (917–1,580) | 273 (115–449) | 1,570 (1,080–2,010) | 1,100 (850–1,410) |
| GAMBIA | NA | NA | NA | 139 (39–234) |
| GUINEA-BISSAU | NA | NA | 80 (21–136) | 68 (22–115) |
| GRENADA | 19 (15–23) | 3 (1–6) | 6 (3–8) | 16 (11–20) |
| GUATEMALA | 395 (215–555) | 1,120 (545–1,670) | 728 (469–967) | 822 (531–1,120) |
| GUYANA | 24 (7–44) | NA | NA | 72 (41–107) |
| HONDURAS | 2,190 (1,520–2,850) | 419 (241–599) | 1,020 (628–1,360) | 1,610 (1,160–2,050) |
| HAITI | 450 (211–680) | 1,600 (946–2,190) | 827 (549–1,130) | 133 (63–208) |
| INDONESIA | 10,100 (7,570–13,300) | 13,500 (8,770–17,800) | 9,080 (6,710–12,100) | 38,400 (29,000–49,700) |
| INDIA | 95,300 (72,100–118,000) | 143,000 (92,600–189,000) | 133,000 (98,500–177,000) | 133,000 (103,000–169,000) |
| IRAN, ISLAMIC REPUBLIC OF | 3,100 (2,730–3,450) | 1,570 (715–2,200) | 1,250 (1,020–1,510) | 1,280 (1,020–1,560) |
| IRAQ | 2,180 (1,670–2,760) | 516 (242–787) | 1,540 (1,060–2,030) | 3,060 (2,420–3,870) |
| JAMAICA | 119 (90–147) | 6 (1–13) | 53 (37–74) | 102 (70–127) |
| JORDAN | 758 (532–988) | 225 (120–350) | 725 (559–939) | 892 (662–1,120) |
| KENYA | 19,000 (13,900–24,900) | 1,900 (1,180–2,660) | 3,990 (2,890–5,420) | 13,600 (10,100–17,600) |
| KYRGYZSTAN | 507 (368–643) | 227 (105–398) | 345 (160–517) | 1,490 (1,010–1,970) |
| CAMBODIA | 757 (498–998) | 196 (82–347) | 295 (218–389) | 2,050 (1,320–2,770) |
| KIRIBATI | 7 (5–9) | 16 (9–22) | 3 (1–6) | 17 (10–24) |
| LAO PEOPLE'S DEMOCRATIC REPUBLIC | 555 (377–700) | 549 (320–782) | 371 (258–506) | 1,160 (877–1,430) |
| LIBERIA | 517 (375–651) | 16 (3–35) | 597 (426–791) | 404 (252–568) |
| ST LUCIA | NA | NA | NA | NA |
| SRI LANKA | 3,490 (2,630–4,490) | 181 (53–323) | 741 (517–970) | 3,190 (2,110–4,100) |
| LESOTHO | 315 (208–430) | NA | 136 (75–206) | 1,910 (1,320–2,530) |
| MOROCCO | 3,820 (2,710–4,760) | 163 (38–280) | 1,440 (1,070–1,840) | 7,070 (5,340–8,880) |
| MOLDOVA, REPUBLIC OF | 124 (90–156) | 14 (4–26) | 24 (14–34) | 601 (411–818) |
| MADAGASCAR | 14,100 (11,400–17,000) | 258 (74–500) | 4,010 (2,970–5,150) | 26,400 (19,800–32,300) |
| MALDIVES | NA | NA | NA | NA |

**CUMULATIVE CASES AVERTED BY COUNTRY ACROSS 2030–2100**

| COUNTRY | ANAL | OROPHARYNGEAL | VAGINAL | VULVAR |
|---|---|---|---|---|
| MARSHALL ISLANDS | NA | NA | NA | 3 (0–5) |
| MACEDONIA (NORTH), THE FORMER YUGOSLAV REPUBLIC OF | 134 (94–172) | 33 (16–54) | 56 (26–86) | 128 (77–181) |
| MALI | 6,400 (4,930–7,670) | 3,180 (1,790–4,550) | 5,800 (4,500–7,220) | 7,900 (6,160–9,960) |
| MYANMAR | 1,970 (1,060–2,830) | 310 (107–521) | 1,140 (662–1,740) | 4,280 (3,100–5,620) |
| MONGOLIA | 116 (53–173) | 34 (10–61) | 329 (234–426) | 359 (260–493) |
| MOZAMBIQUE | 13,000 (9,760–16,700) | 145 (30–346) | 4,370 (2,340–6,480) | 40,700 (31,300–52,600) |
| MAURITANIA | 1,310 (1,020–1,620) | 259 (123–431) | 718 (532–926) | 12,800 (7,520–18,100) |
| MALAWI | 10,200 (7,790–12,500) | NA | 7,890 (5,940–10,100) | 10,200 (7,480–13,100) |
| NAMIBIA | 455 (327–583) | 72 (18–134) | 606 (423–796) | 1,580 (1,190–2,040) |
| NIGER | 7,780 (5,280–10,400) | 1,540 (742–2,530) | 5,430 (4,030–7,150) | 3,430 (2,410–4,470) |
| NIGERIA | 46,800 (34,500–61,300) | 10,400 (3,710–19,100) | 15,500 (9,930–21,900) | 100,000 (74,200–131,000) |
| NICARAGUA | 875 (643–1,150) | 397 (232–556) | 361 (256–476) | 589 (396–784) |
| NEPAL | 786 (520–1,020) | 548 (278–803) | 1,110 (742–1,500) | 946 (635–1,290) |
| PAKISTAN | 19,600 (15,700–24,400) | 21,700 (15,300–29,300) | 17,100 (12,200–22,800) | 21,700 (15,800–28,500) |
| PERU | 1,940 (1,540–2,330) | 2,250 (1,270–2,930) | 1,590 (1,190–2,000) | 2,200 (1,670–2,770) |
| PHILIPPINES | 4,440 (3,330–5,560) | 1,070 (343–1,840) | 3,340 (2,490–4,460) | 8,900 (6,920–11,200) |
| PAPUA NEW GUINEA | 3,380 (2,250–4,460) | 400 (92–745) | 847 (420–1,260) | 985 (623–1,390) |
| KOREA, DEMOCRATIC PEOPLE'S REPUBLIC OF | 391 (292–490) | 189 (100–257) | 391 (255–530) | 889 (652–1,140) |
| PARAGUAY | 2,260 (1,690–2,770) | 361 (164–583) | 39 (11–65) | 1,740 (1,270–2,320) |
| PALESTINE, STATE OF | 526 (402–645) | 334 (189–478) | 284 (206–377) | 973 (686–1,250) |
| RWANDA | 2,310 (1,770–2,870) | 51 (18–92) | 1,100 (798–1,420) | 10,000 (8,440–12,000) |
| SUDAN | 4,700 (2,250–7,170) | 1,590 (923–2,370) | 4,490 (1,310–7,970) | 16,400 (11,300–21,900) |
| SENEGAL | 211 (60–375) | NA | NA | NA |
| SOLOMON ISLANDS | 234 (174–306) | 15 (3–24) | 130 (92–171) | 676 (518–839) |
| SIERRA LEONE | 1,490 (1,140–1,930) | 28 (7–52) | 937 (700–1,220) | 3,020 (2,360–3,720) |
| EL SALVADOR | 395 (293–504) | 67 (30–107) | 90 (56–127) | 279 (201–360) |
| SOMALIA | 4,070 (3,110–5,200) | 149 (37–295) | 1,390 (905–1,850) | 7,270 (5,700–9,040) |
| SERBIA | 818 (302–1,300) | NA | 208 (59–365) | NA |
| SOUTH SUDAN | NA | 502 (118–1,170) | 1,700 (489–3,000) | 4,070 (1,290–7,040) |
| SAO TOME AND PRINCIPE | 22 (15–29) | 12 (4–19) | 13 (9–18) | 53 (38–74) |

**CUMULATIVE CASES AVERTED BY COUNTRY ACROSS 2030–2100**

| COUNTRY | ANAL | OROPHARYNGEAL | VAGINAL | VULVAR |
|---|---|---|---|---|
| SWAZILAND | 587 (383–790) | NA | 242 (133–364) | 1,900 (1,420–2,370) |
| SYRIAN ARAB REPUBLIC | 970 (718–1,290) | 418 (228–636) | 750 (523–955) | 2,790 (1,990–3,730) |
| CHAD | NA | 207 (51–451) | 1,980 (836–3,540) | 5,830 (2,680–8,720) |
| TOGO | 1,030 (807–1,290) | 206 (64–378) | 1,340 (912–1,920) | 2,320 (1,810–2,940) |
| THAILAND | 1,890 (1,480–2,290) | 347 (99–633) | 1,310 (962–1,720) | 3,500 (2,760–4,310) |
| TAJIKISTAN | 1,340 (990–1,710) | 762 (448–1,100) | 52 (16–92) | 768 (527–1,040) |
| TURKMENISTAN | 1,460 (1,150–1,760) | 1,220 (749–1,670) | 460 (326–624) | 1,170 (881–1,520) |
| TIMOR-LESTE | NA | NA | NA | NA |
| TONGA | 3 (1–6) | NA | 4 (1–7) | 7 (4–10) |
| TUNISIA | 1,200 (941–1,460) | 164 (84–249) | 431 (322–550) | 1,470 (1,140–1,800) |
| TUVALU | NA | NA | NA | NA |
| TANZANIA, UNITED REPUBLIC OF | 57,000 (41,800–70,100) | 4,750 (1,330–8,450) | 15,900 (11,700–20,400) | 46,100 (28,400–61,900) |
| UGANDA | 6,130 (4,520–7,880) | 3,560 (1,650–5,470) | 5,340 (3,790–7,050) | 20,300 (13,900–26,800) |
| UKRAINE | 2,030 (1,460–2,490) | 2,000 (1,130–2,720) | 946 (692–1,210) | 4,680 (3,440–6,050) |
| UZBEKISTAN | 8,560 (6,590–10,600) | 4,840 (2,930–6,410) | 1,760 (1,250–2,420) | 4,380 (3,100–5,650) |
| ST VINCENT & THE GRENADINES | NA | NA | NA | NA |
| VENEZUELA, BOLIVARIAN REPUBLIC OF | 5,910 (4,260–7,670) | 1,780 (992–2,490) | 2,540 (1,710–3,370) | 2,070 (1,510–2,680) |
| VIET NAM | 7,550 (5,880–9,290) | 2,820 (1,860–3,720) | 1,030 (774–1,370) | 4,000 (3,050–5,360) |
| VANUATU | 110 (74–147) | NA | NA | NA |
| SAMOA | 26 (20–32) | 1 (0–3) | 18 (11–25) | 37 (29–44) |
| KOSOVO | 139 (108–172) | 10 (3–16) | 61 (44–82) | 400 (303–505) |
| YEMEN | 745 (338–1,170) | 781 (276–1,540) | 3,150 (2,160–4,150) | 2,950 (1,960–3,860) |
| SOUTH AFRICA | 3,760 (2,580–4,940) | 2,490 (1,410–3,460) | 3,090 (2,220–3,970) | 2,570 (1,740–3,520) |
| ZAMBIA | 7,840 (5,660–9,870) | 122 (40–218) | 5,690 (3,690–7,980) | 20,100 (15,100–25,300) |
| ZIMBABWE | 5,610 (4,480–7,070) | 146 (42–273) | 3,160 (2,240–4,250) | 17,900 (13,900–22,400) |

**CUMULATIVE DEATHS AVERTED BY COUNTRY ACROSS 2030–2100**

| COUNTRY | ANAL | OROPHARYNGEAL | VAGINAL | VULVAR |
|---|---|---|---|---|
| AFGHANISTAN | 5,120 (3,930–6,440) | 4,720 (2,830–6,190) | 1,550 (1,130–2,010) | 3,680 (2,630–4,840) |
| ANGOLA | 1,740 (1,150–2,390) | 7,190 (3,940–10,800) | 5,140 (3,250–7,530) | 5,000 (2,990–7,350) |
| ALBANIA | 33 (17–50) | 3 (1–5) | 57 (39–81) | 73 (38–117) |
| ARMENIA | 70 (43–104) | NA | 75 (46–111) | 107 (58–175) |

### CUMULATIVE DEATHS AVERTED BY COUNTRY ACROSS 2030–2100

| COUNTRY | ANAL | OROPHARYNGEAL | VAGINAL | VULVAR |
|---|---|---|---|---|
| **AZERBAIJAN** | 529 (309–784) | 32 (7–61) | 28 (13–44) | 394 (243–537) |
| **BURUNDI** | 2,210 (1,610–2,840) | 98 (26–194) | 1,070 (750–1,450) | 4,380 (2,880–6,150) |
| **BENIN** | 1,370 (1,020–1,700) | 1,500 (745–2,080) | 1,190 (868–1,500) | 2,320 (1,690–3,080) |
| **BURKINA FASO** | 9,910 (6,410–13,600) | NA | 1,840 (491–3,370) | 2,990 (1,590–4,550) |
| **BANGLADESH** | 3,500 (2,600–4,450) | 2,220 (707–4,070) | 4,560 (3,300–6,090) | 9,400 (6,920–12,000) |
| **BOSNIA AND HERZEGOVINA** | 41 (24–60) | 65 (32–100) | 54 (37–72) | 45 (13–87) |
| **BELARUS** | 430 (302–559) | 42 (12–76) | 125 (87–168) | 276 (79–543) |
| **BELIZE** | NA | NA | NA | NA |
| **BOLIVIA, PLURINATIONAL STATE OF** | 326 (201–472) | 1,270 (407–2,580) | 991 (525–1,460) | 1,010 (533–1,440) |
| **BHUTAN** | NA | 213 (120–338) | NA | NA |
| **CENTRAL AFRICAN REPUBLIC** | 878 (621–1,150) | NA | 293 (79–511) | 1,370 (925–1,900) |
| **CHINA** | 14,300 (11,300–18,200) | 9,140 (4,120–13,500) | 13,500 (9,810–17,600) | 20,500 (15,200–27,300) |
| **COTE D'IVOIRE** | 4,730 (3,410–5,980) | 1,690 (850–2,650) | 3,970 (2,650–5,480) | 2,980 (2,000–4,080) |
| **CAMEROON** | 3,430 (2,410–4,460) | 333 (108–614) | 2,250 (1,510–2,980) | 3,860 (2,730–5,230) |
| **CONGO, THE DEMOCRATIC REPUBLIC OF THE** | 26,100 (20,100–32,600) | 325 (91–587) | 8,600 (5,950–11,300) | 34,600 (24,500–46,600) |
| **CONGO** | 939 (649–1,240) | 155 (49–280) | 314 (211–425) | 1,190 (832–1,640) |
| **COLOMBIA** | 6,640 (5,790–7,570) | 475 (134–797) | 2,450 (1,970–2,920) | 5,790 (4,040–7,710) |
| **COMOROS** | NA | NA | NA | 178 (112–265) |
| **CABO VERDE (CAPE VERDE)** | 30 (7–50) | NA | NA | 23 (6–41) |
| **CUBA** | 664 (450–858) | 40 (12–75) | 221 (151–298) | 513 (352–727) |
| **DJIBOUTI** | NA | NA | 43 (21–66) | 123 (82–167) |
| **DOMINICA** | NA | 1 (0–3) | 6 (2–10) | 1 (0–2) |
| **ALGERIA** | 1,870 (1,370–2,340) | 513 (303–702) | 1,120 (853–1,420) | 2,210 (1,490–3,040) |
| **ECUADOR** | 3,050 (1,990–4,170) | 185 (64–330) | 768 (542–1,020) | 2,550 (1,810–3,430) |
| **EGYPT** | 4,620 (3,600–5,740) | 696 (266–1,260) | 2,880 (2,070–3,810) | 8,710 (6,130–11,500) |

**CUMULATIVE DEATHS AVERTED BY COUNTRY ACROSS 2030–2100**

| COUNTRY | ANAL | OROPHARYNGEAL | VAGINAL | VULVAR |
|---|---|---|---|---|
| ERITREA | 263 (152–393) | NA | 277 (168–407) | 632 (450–910) |
| ETHIOPIA | 19,900 (14,300–25,300) | 3,420 (2,000–5,070) | 17,600 (12,800–22,900) | 35,400 (24,800–46,900) |
| FIJI | 204 (116–282) | 7 (1–14) | 47 (20–73) | NA |
| MICRONESIA, FEDERATED STATES OF | NA | NA | NA | NA |
| GEORGIA | 43 (25–59) | 8 (3–16) | 61 (41–84) | 168 (69–283) |
| GHANA | 5,550 (3,520–7,860) | 232 (55–516) | 994 (606–1,480) | 4,200 (2,970–5,580) |
| GUINEA | 734 (539–964) | 210 (89–356) | 1,160 (790–1,520) | 554 (362–779) |
| GAMBIA | NA | NA | NA | 113 (32–194) |
| GUINEA-BISSAU | NA | NA | 52 (14–90) | 37 (12–66) |
| GRENADA | 12 (8–16) | 2 (0–4) | 4 (2–6) | 10 (5–14) |
| GUATEMALA | 312 (167–431) | 790 (384–1,220) | 554 (344–761) | 598 (381–857) |
| GUYANA | 20 (5–35) | NA | NA | 53 (28–83) |
| HONDURAS | 1,360 (925–1,930) | 291 (171–422) | 754 (465–1,020) | 1,090 (766–1,440) |
| HAITI | 416 (184–635) | 781 (449–1,130) | 568 (377–784) | 98 (48–155) |
| INDONESIA | 6,490 (4,790–8,730) | 9,310 (5,810–12,600) | 5,910 (4,320–8,250) | 21,900 (16,500–28,400) |
| INDIA | 65,900 (51,200–84,000) | 87,000 (54,700–117,000) | 73,000 (52,400–98,900) | 81,200 (60,000–104,000) |
| IRAN, ISLAMIC REPUBLIC OF | 2,070 (1,820–2,350) | 973 (445–1,390) | 914 (726–1,120) | 873 (665–1,110) |
| IRAQ | 1,770 (1,350–2,220) | 404 (189–607) | 1,190 (814–1,540) | 2,170 (1,710–2,760) |
| JAMAICA | 77 (57–100) | 5 (1–11) | 41 (28–59) | 72 (49–97) |
| JORDAN | 586 (422–770) | 174 (91–273) | 507 (373–669) | 608 (439–799) |
| KENYA | 10,700 (7,600–13,900) | 1,440 (887–2,070) | 2,860 (2,120–3,720) | 7,870 (5,670–10,900) |
| KYRGYZSTAN | 326 (226–430) | 150 (74–253) | 249 (120–377) | 730 (483–1,110) |
| CAMBODIA | 488 (316–674) | 158 (65–284) | 219 (155–284) | 1,490 (922–2,020) |
| KIRIBATI | 4 (3–6) | 12 (7–19) | 1 (0–4) | 6 (2–9) |
| LAO PEOPLE'S DEMOCRATIC REPUBLIC | 395 (270–493) | 457 (273–640) | 272 (182–363) | 751 (539–994) |
| LIBERIA | 356 (257–446) | 13 (3–30) | 384 (275–507) | 287 (181–408) |
| ST LUCIA | NA | NA | NA | NA |
| SRI LANKA | 2,280 (1,610–3,050) | 129 (37–231) | 533 (370–702) | 1,760 (1,030–2,430) |
| LESOTHO | 240 (160–327) | NA | 103 (55–154) | 359 (99–639) |

**CUMULATIVE DEATHS AVERTED BY COUNTRY ACROSS 2030–2100**

| COUNTRY | ANAL | OROPHARYNGEAL | VAGINAL | VULVAR |
|---|---|---|---|---|
| **MOROCCO** | 2,120 (1,460–2,700) | 121 (29–204) | 983 (741–1,280) | 2,930 (2,090–3,810) |
| **MOLDOVA, REPUBLIC OF** | 94 (67–119) | 9 (3–16) | 17 (10–25) | 138 (48–246) |
| **MADAGASCAR** | 5,830 (4,380–7,460) | 206 (59–406) | 2,650 (1,900–3,480) | 10,300 (6,280–15,100) |
| **MALDIVES** | NA | NA | NA | NA |
| **MARSHALL ISLANDS** | NA | NA | NA | NA |
| **MACEDONIA (NORTH), THE FORMER YUGOSLAV REPUBLIC OF** | 106 (74–134) | 24 (12–38) | 41 (19–66) | 55 (28–95) |
| **MALI** | 4,280 (3,240–5,380) | 2,430 (1,350–3,410) | 4,010 (3,090–5,030) | 5,020 (3,600–6,690) |
| **MYANMAR** | 1,490 (743–2,160) | 240 (81–402) | 762 (431–1,180) | 2,880 (2,080–3,810) |
| **MONGOLIA** | 84 (43–127) | 21 (6–39) | 183 (127–249) | 211 (131–301) |
| **MOZAMBIQUE** | 6,480 (4,650–8,620) | 127 (26–309) | 3,290 (1,840–4,850) | 14,800 (8,570–22,700) |
| **MAURITANIA** | 739 (577–975) | 205 (99–343) | 488 (362–636) | 7,890 (4,590–11,600) |
| **MALAWI** | 4,440 (3,340–5,760) | NA | 3,840 (2,840–5,070) | 4,810 (3,140–6,810) |
| **NAMIBIA** | 302 (208–402) | 43 (10–82) | 278 (195–370) | 543 (315–800) |
| **NIGER** | 5,200 (3,430–7,050) | 1,700 (782–2,610) | 3,760 (2,780–5,010) | 1,820 (1,200–2,540) |
| **NIGERIA** | 28,100 (20,700–36,700) | 8,460 (2,880–15,400) | 11,900 (7,510–17,300) | 47,900 (30,700–66,600) |
| **NICARAGUA** | 522 (373–688) | 277 (156–394) | 206 (139–280) | 353 (206–519) |
| **NEPAL** | 628 (421–838) | 391 (202–584) | 663 (460–892) | 729 (490–1,010) |
| **PAKISTAN** | 15,300 (12,000–19,200) | 15,600 (10,800–21,100) | 12,900 (9,260–17,200) | 15,900 (11,300–21,400) |
| **PERU** | 1,190 (902–1,500) | 1,630 (917–2,250) | 1,160 (861–1,500) | 1,220 (836–1,680) |
| **PHILIPPINES** | 2,930 (2,110–3,740) | 733 (232–1,330) | 2,180 (1,470–2,900) | 5,900 (4,440–7,570) |
| **PAPUA NEW GUINEA** | 1,710 (1,100–2,340) | 297 (69–554) | 511 (244–781) | 739 (463–1,050) |
| **KOREA, DEMOCRATIC PEOPLE'S REPUBLIC OF** | 233 (170–300) | 129 (68–183) | 285 (182–409) | 616 (448–811) |
| **PARAGUAY** | 1,450 (1,050–1,770) | 273 (128–444) | 30 (9–50) | 981 (671–1,480) |

**CUMULATIVE DEATHS AVERTED BY COUNTRY ACROSS 2030–2100**

| COUNTRY | ANAL | OROPHARYNGEAL | VAGINAL | VULVAR |
|---|---|---|---|---|
| PALESTINE, STATE OF | 365 (284–446) | 274 (154–395) | 206 (149–271) | 549 (392–733) |
| RWANDA | 1,650 (1,220–2,110) | 38 (13–67) | 839 (617–1,080) | 4,610 (3,400–6,210) |
| SUDAN | 3,770 (1,850–5,790) | 1,060 (609–1,600) | 3,380 (1,010–5,890) | 8,930 (5,900–12,200) |
| SENEGAL | 152 (44–273) | NA | NA | NA |
| SOLOMON ISLANDS | 162 (113–212) | 11 (2–18) | 91 (63–123) | 346 (233–478) |
| SIERRA LEONE | 1,010 (745–1,310) | 22 (6–39) | 639 (473–837) | 1,750 (1,270–2,250) |
| EL SALVADOR | 274 (199–346) | 45 (20–73) | 51 (29–74) | 118 (71–172) |
| SOMALIA | 2,570 (1,840–3,310) | 116 (31–230) | 939 (631–1,290) | 3,880 (2,740–5,240) |
| SERBIA | 660 (252–1,030) | NA | 190 (54–339) | NA |
| SOUTH SUDAN | NA | 414 (99–960) | 1,300 (379–2,280) | 3,210 (1,030–5,580) |
| SAO TOME AND PRINCIPE | 11 (7–15) | 6 (2–11) | 6 (3–9) | 7 (1–13) |
| SWAZILAND | 438 (277–601) | NA | 182 (99–265) | 251 (67–452) |
| SYRIAN ARAB REPUBLIC | 689 (495–934) | 297 (165–455) | 525 (354–700) | 1,550 (1,050–2,180) |
| CHAD | NA | 185 (44–404) | 1,400 (565–2,400) | 3,800 (1,550–5,900) |
| TOGO | 662 (497–847) | 168 (52–299) | 908 (606–1,310) | 1,230 (849–1,610) |
| THAILAND | 1,230 (955–1,550) | 233 (71–422) | 856 (625–1,100) | 2,190 (1,610–2,760) |
| TAJIKISTAN | 1,010 (722–1,370) | 488 (289–725) | 41 (13–72) | 518 (358–723) |
| TURKMENISTAN | 878 (643–1,130) | 651 (397–957) | 334 (223–468) | 695 (452–982) |
| TIMOR-LESTE | NA | NA | NA | NA |
| TONGA | 2 (1–4) | NA | 2 (1–5) | 4 (2–7) |
| TUNISIA | 709 (558–861) | 110 (54–167) | 315 (236–406) | 772 (566–1,010) |
| TUVALU | NA | NA | NA | NA |
| TANZANIA, UNITED REPUBLIC OF | 21,400 (15,200–28,300) | 3,800 (1,080–6,830) | 9,660 (7,040–12,500) | 22,500 (14,500–33,000) |
| UGANDA | 4,540 (3,190–5,820) | 2,690 (1,230–4,130) | 3,980 (2,770–5,290) | 13,000 (8,500–18,300) |
| UKRAINE | 1,130 (751–1,450) | 1,210 (675–1,710) | 558 (396–712) | 874 (273–1,540) |
| UZBEKISTAN | 4,750 (3,410–6,280) | 2,940 (1,840–3,890) | 1,290 (921–1,750) | 2,970 (1,960–4,080) |
| ST VINCENT & THE GRENADINES | NA | NA | NA | NA |

CUMULATIVE DEATHS AVERTED BY COUNTRY ACROSS 2030–2100

| COUNTRY | ANAL | OROPHARYNGEAL | VAGINAL | VULVAR |
|---|---|---|---|---|
| **VENEZUELA, BOLIVARIAN REPUBLIC OF** | 2,920 (1,940–4,010) | 1,170 (675–1,660) | 1,530 (1,030–2,020) | 1,180 (793–1,580) |
| **VIET NAM** | 4,170 (3,230–5,220) | 2,150 (1,360–2,840) | 777 (564–1,050) | 2,790 (1,990–3,740) |
| **VANUATU** | 92 (61–125) | NA | NA | NA |
| **SAMOA** | 17 (13–21) | 1 (0–2) | 12 (7–18) | 22 (17–29) |
| **KOSOVO** | 89 (68–113) | 6 (2–11) | 44 (30–60) | 141 (71–217) |
| **YEMEN** | 640 (297–992) | 547 (189–1,070) | 2,290 (1,570–3,120) | 1,770 (1,170–2,350) |
| **SOUTH AFRICA** | 2,390 (1,610–3,160) | 1,630 (914–2,330) | 2,060 (1,460–2,830) | 1,450 (967–1,990) |
| **ZAMBIA** | 4,950 (3,660–6,270) | 88 (28–159) | 3,720 (2,430–5,320) | 7,750 (4,940–11,200) |
| **ZIMBABWE** | 2,690 (1,990–3,610) | 112 (33–203) | 2,020 (1,370–2,770) | 6,050 (3,880–8,860) |

**Table S7: Country-wise impact of HPV vaccination on cancer outcomes (2030–2100).**

Note: 95% uncertainty interval in parentheses

lb= lower bound; NA= not applicable; ub= upper bound.

| Country Name | Region | Income Level | Probability of Death for Anal Cancer (lb, ub) | Probability of Death for Oropharyngeal Cancer (lb, ub) | Probability of Death for Vaginal Cancer (lb, ub) | Probability of Death for Vulvar Cancer (lb, ub) | Source |
|---|---|---|---|---|---|---|---|
| Afghanistan | EMR | low | 0.88 (0.79, 0.97) * | 0.87 (0.81, 0.93) | 0.92 (0.87, 0.97) | 0.88 (0.75, 1) | 19 |
| Angola | AFR | lmi | 0.76 (0.64, 0.87) | 0.93 (0.84, 1) | 0.74 (0.65, 0.83) | 0.55 (0.36, 0.74) | 19 |
| Albania | EUR | umi | 1 (0.85, 1) | 0.57 (0.45, 0.68) | 1 (0.88, 1) * | 0.43 (0.17, 0.68) | 19 |
| Armenia | EUR | umi | 1 (0.85, 1) * | 1 (0.88, 1) * | 0.94 (0.82, 1) | 0.64 (0.38, 0.89) | 19 |
| Azerbaijan | EUR | umi | 0.65 (0.5, 0.8) | 0.77 (0.65, 0.89) | 0.96 (0.84, 1) | 0.85 (0.6, 1) | 19 |
| Burundi | AFR | low | 0.7 (0.59, 0.82) | 0.95 (0.86, 1) | 0.88 (0.79, 0.96) | 0.59 (0.4, 0.79) | 19 |
| Benin | AFR | lmi | 0.88 (0.77, 0.99) | 0.82 (0.73, 0.91) | 0.89 (0.8, 0.97) | 0.76 (0.56, 0.95) | 19 |
| Burkina Faso | AFR | low | 0.86 (0.75, 0.97) | 0.91 (0.82, 1) | 0.77 (0.68, 0.86) | 0.7 (0.51, 0.89) | 19 |
| Bangladesh | SEAR | lmi | 0.93 (0.85, 1) | 0.64 (0.52, 0.75) | 0.86 (0.76, 0.96) | 0.84 (0.74, 0.95) | 19 |
| Bosnia and Herzegovina | EUR | umi | 0.84 (0.68, 0.99) | 0.79 (0.67, 0.9) | 0.7 (0.58, 0.81) | 0.24 (0, 0.49) | 19 |
| Belarus | EUR | umi | 0.55 (0.4, 0.71) | 0.71 (0.6, 0.83) | 0.84 (0.72, 0.95) | 0.21 (0, 0.46) | 19 |
| Belize | AMR | umi | 1 (0.87, 1) * | 1 (0.88, 1) * | 1 (0.91, 1) * | 1 (0.77, 1)* | 19 |
| Bolivia, Plurinational State of | AMR | lmi | 1 (0.87, 1) * | 0.81 (0.69, 0.93) | 0.83 (0.74, 0.91) | 0.86 (0.63, 1) | 19 |
| Bhutan | SEAR | lmi | 1 (0.92, 1) * | 0.76 (0.64, 0.87) | 0.76 (0.66, 0.86) | 1 (0.89, 1) * | 19 |
| Central African Republic | AFR | low | 0.75 (0.64, 0.86) | 0.96 (0.87, 1) | 0.89 (0.8, 0.97) | 0.69 (0.5, 0.88) | 19 |
| China (Asia) | WPR | umi | 0.86 (0.75, 0.97) | 0.93 (0.86, 1) | 0.9 (0.76, 1) | 0.84 (0.66, 1) | 19 |
| Cote d'Ivoire | AFR | lmi | 0.76 (0.64, 0.87) | 0.92 (0.83, 1.01) | 0.8 (0.72, 0.89) | 0.87 (0.68, 1) | 19 |
| Cameroon | AFR | lmi | 0.77 (0.66, 0.11) | 0.87 (0.78, 0.96) | 0.81 (0.72, 0.9) | 0.79 (0.6, 0.99) | 19 |
| Congo, the Democratic Republic of the | AFR | low | 0.76 (0.64, 0.87) | 0.95 (0.86, 1) | 0.82 (0.73, 0.91) | 0.66 (0.46, 0.85) | 19 |

| | | | | | | | |
|---|---|---|---|---|---|---|---|
| Congo | AFR | lmi | 0.65 (0.54, 0.76) | 1 (0.91, 1) * | 0.9 (0.81, 0.98) | 0.7 (0.51, 0.89) | 19 (assumption for survival rate of all cancers based on AFR) |
| Colombia | AMR | umi | 0.78 (0.65, 0.91) | 0.79 (0.68, 0.91) | 0.84 (0.75, 0.92) | 0.59 (0.36, 0.83) | 19 |
| Comoros | AFR | lmi | 0.79 (0.68, 0.91) | 1 (0.91, 1) * | 1 (0.91, 1) * | 0.39 (0.2, 0.59) | 19 |
| Cabo Verde | AFR | lmi | 0.79 (0.68, 0.91) | 0.49 (0.4, 0.58) | 1 (0.91, 1) * | 0.7 (0.51, 0.89) | 19 |
| Cuba | AMR | umi | 0.56 (0.43, 0.69) | 0.58 (0.47, 0.7) | 0.67 (0.58, 0.76) | 0.5 (0.26, 0.73) | 19 |
| Djibouti | EMR | lmi | 1 (0.91, 1) * | 1 (0.94, 1) * | 1 (0.95, 1) * | 0.81 (0.68, 0.94) | 19 |
| Dominica | AMR | umi | 0.82 (0.69, 0.95) | 0.86 (0.74, 0.98) | 0.84 (0.76, 0.93) | 0.7 (0.46, 0.93) | 19 (assumption for survival rate of anal, and oropharyngeal cancers based on AMR) |
| Algeria | AFR | lmi | 0.9 (0.79, 1) | 0.96 (0.87, 1) | 0.92 (0.84, 1) | 0.86 (0.67, 1) | 19 |
| Ecuador | AMR | umi | 0.8 (0.68, 0.93) | 0.87 (0.75, 0.99) | 0.9 (0.81, 0.98) | 0.76 (0.52, 0.99) | 19 |
| Egypt | EMR | lmi | 0.91 (0.82, 1) | 0.93 (0.88, 0.99) | 0.93 (0.88, 0.98) | 0.8 (0.67, 0.93) | 19 |
| Eritrea | AFR | low | 0.9 (0.78, 1.01) | 1 (0.91, 1) * | 0.84 (0.76, 0.93) | 0.6 (0.41, 0.79) | 19 |
| Ethiopia | AFR | low | 0.81 (0.7, 0.92) | 0.97 (0.88, 1) | 0.79 (0.7, 0.87) | 0.67 (0.48, 0.86) | 19 |

| | | | | | | | |
|---|---|---|---|---|---|---|---|
| Fiji | WPR | umi | 1 (0.89, 1) * | 1 (0.93, 1) * | 0.79 (0.65, 0.94) | 1 (0.81, 1) * | 19 |
| Micronesia, Federated States of | WPR | lmi | 0.85 (0.74, 0.96) | 0.88 (0.81, 0.95) | 0.78 (0.63, 0.92) | 0.34 (0.15, 0.53) | 19 (assumption for survival rate of anal cancer based on WPR) |
| Georgia | EUR | umi | 0.9 (0.75, 1.06) | 0.9 (0.79, 1) | 0.68 (0.57, 0.8) | 0.28 (0.02, 0.53) | 19 |
| Ghana | AFR | lmi | 0.64 (0.53, 0.76) | 0.9 (0.81, 1) | 0.93 (0.85, 1) | 0.82 (0.63, 1) | 19 |
| Guinea | AFR | lmi | 0.73 (0.62, 0.85) | 0.9 (0.81, 1) | 0.86 (0.77, 0.95) | 0.63 (0.43, 0.82) | 19 (assumption for survival rate of all cancers based on AFR) |
| Gambia | AFR | low | 0.92 (0.81, 1) | 1 (0.91, 1) * | 1 (0.91, 1) * | 1 (0.81, 1) * | 19 |
| Guinea-Bissau | AFR | low | 0.9 (0.79, 1.02) | 1 (0.91, 1) * | 0.82 (0.73, 0.91) | 0.82 (0.63, 1) | 19 |
| Grenada | AMR | umi | 0.82 (0.69, 0.95) | 0.86 (0.74, 0.98) | 0.85 (0.77, 0.94) | 0.71 (0.48, 0.95) | 19 (assumption for survival rate of anal and oropharyngeal cancers based on AMR) |
| Guatemala | AMR | umi | 0.99 (0.86, 1) | 0.88 (0.76, 1) | 0.93 (0.85, 1) | 0.93 (0.7, 1) | 19 |
| Guyana | AMR | | 1 (0.87, 1) * | 1 (0.88, 1) * | 1 (0.91, 1) * | 1 (0.77, 1) * | 19 |

| | | | | | | | |
|---|---|---|---|---|---|---|---|
| Honduras | AMR | lmi | 0.74 (0.61, 0.87) | 0.85 (0.73, 0.97) | 0.85 (0.77, 0.94) | 0.89 (0.65, 1) | 19 |
| Haiti | AMR | lmi | 0.98 (0.85, 1) | 0.65 (0.53, 0.77) | 0.83 (0.74, 0.91) | 0.97 (0.74, 1) | 19 |
| Indonesia | SEAR | umi | 0.92 (0.84, 1) | 0.89 (0.77, 1) | 0.85 (0.75, 0.95) | 0.72 (0.61, 0.82) | 19 |
| India | SEAR | lmi | 0.84 (0.75, 0.92) | 0.73 (0.62, 0.85) | 0.66 (0.56, 0.76) | 0.77 (0.67, 0.88) | 19 |
| Iran, Islamic Republic of | EMR | lmi | 0.93 (0.84, 1) | 0.91 (0.86, 0.97) | 0.97 (0.92, 1) | 0.97 (0.84, 1) | 19 |
| Iraq | EMR | umi | 0.95 (0.86, 1) | 0.98 (0.92, 1) | 0.95 (0.9, 1) | 0.93 (0.8, 1) | 19 |
| Jamaica | AMR | umi | 0.82 (0.69, 0.95) | 1 (0.88, 1) * | 0.88 (0.79, 0.96) | 0.82 (0.59, 1) | 19 |
| Jordan | EMR | lmi | 0.94 (0.85, 1) | 0.98 (0.92, 1) | 0.87 (0.82, 0.92) | 0.9 (0.78, 1) | 19 |
| Kenya | AFR | lmi | 0.69 (0.58, 0.8) | 0.96 (0.87, 1) | 0.88 (0.79, 0.96) | 0.72 (0.53, 0.91) | 19 |
| Kyrgyzstan | EUR | | 0.86 (0.71, 1) | 0.89 (0.77, 1) | 0.91 (0.8, 1) | 0.64 (0.38, 0.89) | 19 |
| Cambodia | WPR | lmi | 0.91 (0.8, 1) | 0.85 (0.78, 0.92) | 0.93 (0.79, 1) | 0.84 (0.65, 1) | 19 |
| Kiribati | WPR | lmi | 0.85 (0.74, 0.96) | 0.88 (0.81, 0.95) | 0.8 (0.66, 0.95) | 0.34 (0.15, 0.53) | 19 (assumption for survival rate for vaginal and vulvar cancer based on WPR) |
| Lao People's Democratic Republic | WPR | lmi | 0.9 (0.78, 1) | 0.9 (0.83, 0.96) | 0.84 (0.7, 0.99) | 0.78 (0.59, 0.97) | 19 |
| Liberia | AFR | low | 0.82 (0.71, 0.93) | 0.96 (0.87, 1) | 0.76 (0.67, 0.84) | 0.89 (0.69, 1) | 19 |
| St. Lucia | AMR | umi | 1 (0.87, 1) * | 1 (0.88, 1) * | 1 (0.91, 1) * | 1 (0.77, 1) * | 19 |
| Sri Lanka | SEAR | lmi | 0.81 (0.73, 0.89) | 0.85 (0.74, 0.97) | 0.79 (0.69, 0.89) | 0.82 (0.71, 0.92) | 19 |
| Lesotho | AFR | lmi | 0.84 (0.72, 0.95) | 1 (0.91, 1) * | 0.84 (0.75, 0.92) | 0.21 (0.02, 0.4) | 19 |

| | | | | | | | |
|---|---|---|---|---|---|---|---|
| Morocco | EMR | lmi | 0.69 (0.6, 0.78) | 0.9 (0.85, 0.96) | 0.81 (0.76, 0.86) | 0.56 (0.43, 0.69) | 19 |
| Moldova, Republic of | EUR | umi | 0.87 (0.72, 1) | 0.79 (0.67, 0.9) | 0.87 (0.75, 0.99) | 0.29 (0.03, 0.54) | 19 |
| Madagascar | AFR | low | 0.51 (0.4, 0.62) | 0.94 (0.85, 1) | 0.8 (0.72, 0.89) | 0.44 (0.25, 0.64) | 19 |
| Maldives | SEAR | umi | 1 (0.92, 1) * | 1 (0.89, 1) * | 1 (0.9, 1) * | 1 (0.89, 1) * | 19 |
| Marshall Islands | WPR | umi | 0.93 (0.82, 1) | 1 (0.93, 1) * | 1 (0.85, 1) * | 1 (0.81, 1) * | 19 |
| Macedonia, the former Yugoslav Republic of | EUR | | 1 (0.85, 1) * | 0.9 (0.79, 1) | 0.83 (0.71, 0.94) | 0.51 (0.26, 0.77) | 19 |
| Mali | AFR | low | 0.8 (0.69, 0.92) | 0.89 (0.8, 0.98) | 0.82 (0.73, 0.91) | 0.77 (0.58, 0.96) | 19 |
| Myanmar | SEAR | lmi | 0.9 (0.81, 0.98) | 0.87 (0.76, 0.98) | 0.86 (0.76, 0.96) | 0.83 (0.72, 0.93) | 19 |
| Mongolia | WPR | lmi | 0.89 (0.78, 1) | 0.79 (0.72, 0.85) | 0.74 (0.59, 0.89) | 0.79 (0.6, 0.97) | 19 |
| Mozambique | AFR | low | 0.61 (0.5, 0.73) | 0.98 (0.89, 1) | 0.84 (0.76, 0.93) | 0.42 (0.23, 0.61) | 19 |
| Mauritania | AFR | lmi | 0.7 (0.59, 0.82) | 0.96 (0.87, 1) | 0.8 (0.72, 0.89) | 0.7 (0.51, 0.9) | 19 |
| Malawi | AFR | low | 0.53 (0.42, 0.65) | 1 (0.91, 1) * | 0.58 (0.5, 0.67) | 0.54 (0.35, 0.73) | 19 |
| Namibia | AFR | umi | 0.79 (0.68, 0.91) | 0.73 (0.64, 0.82) | 0.54 (0.46, 0.63) | 0.4 (0.21, 0.59) | 19 |
| Niger | AFR | low | 0.76 (0.64, 0.87) | 0.95 (0.86, 1) | 0.82 (0.73, 0.91) | 0.66 (0.46, 0.85) | 19 (assumption for survival rate of all cancers based on AFR) |
| Nigeria | AFR | lmi | 0.76 (0.65, 0.88) | 0.93 (0.84, 1) | 0.91 (0.83, 1) | 0.59 (0.4, 0.79) | 19 |
| Nicaragua | AMR | lmi | 0.81 (0.68, 0.94) | 1 (0.88, 1) * | 0.81 (0.72, 0.9) | 0.79 (0.55, 1) | 19 |
| Nepal | SEAR | lmi | 0.94 (0.86, 1) | 0.93 (0.82, 1) | 0.89 (0.79, 0.99) | 0.94 (0.84, 1) | 19 |
| Pakistan | EMR | lmi | 0.93 (0.84, 1) | 0.88 (0.82, 0.94) | 0.88 (0.83, 0.93) | 0.9 (0.77, 1) | 19 |

| | | | | | | | |
|---|---|---|---|---|---|---|---|
| Peru | AMR | umi | 0.73 (0.61, 0.86) | 0.9 (0.79, 1) | 0.84 (0.76, 0.93) | 0.66 (0.43, 0.9) | 19 |
| Philippines (Asia) | WPR | lmi | 0.84 (0.73, 0.96) | 0.9 (0.84, 0.97) | 0.83 (0.68, 0.97) | 0.88 (0.69, 1) | 19 (assumption for survival rate of all cancers based on WPR) |
| Papua New Guinea | WPR | lmi | 0.62 (0.51, 0.73) | 0.91 (0.85, 0.98) | 0.71 (0.57, 0.86) | 0.87 (0.68, 1) | 19 |
| Korea, Democratic People's Republic of | SEAR | low | 0.79 (0.71, 0.88) | 0.93 (0.82, 1) | 0.89 (0.79, 0.99) | 0.85 (0.75, 0.96) | 19 |
| Paraguay | AMR | umi | 0.77 (0.64, 0.9) | 0.94 (0.82, 1) | 1 (0.91, 1) * | 0.71 (0.48, 0.95) | 19 |
| Palestine, State of | EMR | lmi | 0.89 (0.8, 0.98) | 0.93 (0.88, 0.99) | 0.89 (0.84, 0.94) | 0.79 (0.66, 0.92) | 19 (assumption for survival rate of all cancers based on EMR) |
| Rwanda | AFR | low | 0.84 (0.72, 0.95) | 0.84 (0.75, 0.93) | 0.9 (0.81, 0.98) | 0.54 (0.35, 0.74) | 19 |
| Sudan | EMR | low | 0.9 (0.82, 0.99) | 0.97 (0.91, 1) | 0.9 (0.81, 0.96) | 0.7 (0.57, 0.83) | 19 |
| Senegal | AFR | lmi | 0.76 (0.64, 0.87) | 0.94 (0.85, 1) | 0.82 (0.73, 0.91) | 0.76 (0.57, 0.96) | 19 |
| Solomon Islands | WPR | lmi | 1 (0.89, 1) * | 1 (0.93, 1) * | 1 (0.85, 1) * | 1 (0.81, 1) * | 19 |
| Sierra Leone | AFR | low | 0.79 (0.68, 0.91) | 0.95 (0.86, 1) | 0.82 (0.73, 0.91) | 0.74 (0.55, 0.93) | 19 |
| El Salvador | AMR | umi | 0.84 (0.72, 0.97) | 1 (0.88, 1) * | 0.87 (0.78, 0.96) | 0.65 (0.42, 0.88) | 19 |
| Somalia | EMR | low | 0.79 (0.71, 0.88) | 0.96 (0.9, 1) | 0.81 (0.76, 0.86) | 0.68 (0.55, 0.81) | 19 |

| | | | | | | | |
|---|---|---|---|---|---|---|---|
| Serbia | EUR | umi | 0.67 (0.52, 0.82) | 0.68 (0.57, 0.8) | 0.64 (0.52, 0.75) | 0.14 (0, 0.39) | 19 |
| South Sudan | AFR | low | 0.76 (0.64, 0.87) | 0.95 (0.86, 1) | 0.82 (0.73, 0.91) | 0.66 (0.46, 0.85) | 19 (assumption for survival rate of all cancers based on AFR) |
| Sao Tome and Principe | AFR | lmi | 1 (0.89, 1) * | 1 (0.91, 1) * | 1 (0.91, 1) * | 1 (0.81, 1) | 19 |
| Swaziland | AFR | | 0.84 (0.73, 0.96) | 1 (0.91, 1) * | 0.84 (0.76, 0.93) | 0.13 (0, 0.32) | 19 |
| Syrian Arab Republic | EMR | low | 0.94 (0.85, 1) | 0.94 (0.88, 1) | 0.9 (0.76, 0.95) | 0.74 (0.61, 0.87) | 19 |
| Chad | AFR | low | 0.83 (0.71, 0.94) | 0.94 (0.85, 1) | 0.88 (0.79, 0.96) | 0.76 (0.57, 0.96) | 19 |
| Togo | AFR | low | 0.76 (0.64, 0.87) | 0.95 (0.86, 1) | 0.82 (0.73, 0.91) | 0.66 (0.47, 0.86) | 19 (assumption for survival rate of anal, oropharyngeal, and vaginal cancers based on AFR) |
| Thailand | SEAR | umi | 0.84 (0.75, 0.92) | 0.8 (0.69, 0.92) | 0.79 (0.7, 0.89) | 0.78 (0.67, 0.88) | 19 |
| Tajikistan | EUR | lmi | 0.9 (0.75, 1) | 0.84 (0.73, 0.96) | 1 (0.88, 1) * | 0.92 (0.67, 1) | 19 |
| Turkmenistan | EUR | umi | 0.72 (0.57, 0.87) | 0.63 (0.51, 0.75) | 0.88 (0.76, 1) | 0.72 (0.46, 0.97) | 19 |
| Timor-Leste | SEAR | lmi | 1 (0.92, 1) * | 1 (0.89, 1) * | 1 (0.9, 1) * | 1 (0.89, 1) * | 19 |
| Tonga | WPR | umi | 0.93 (0.82, 1) | 0.98 (0.91, 1) | 0.85 (0.71, 1) | 0.84 (0.66, 1) | 19 |

| | | | | | | | |
|---|---|---|---|---|---|---|---|
| Tunisia | EMR | lmi | 0.75 (0.66, 0.84) | 0.97 (0.91, 1) | 0.9 (0.85, 0.96) | 0.73 (0.6, 0.86) | 19 |
| Tuvalu | WPR | umi | 0.93 (0.82, 1) | 0.98 (0.91, 1) | 0.85 (0.71, 1) | 0.84 (0.66, 1) | 19 |
| Tanzania, United Republic of | AFR | lmi | 0.46 (0.35, 0.57) | 0.94 (0.85, 1) | 0.77 (0.68, 0.86) | 0.55 (0.36, 0.75) | 19 |
| Uganda | AFR | low | 0.88 (0.77, 0.99) | 0.96 (0.87, 1) | 0.9 (0.82, 0.99) | 0.73 (0.54, 0.93) | 19 |
| Ukraine | EUR | lmi | 0.66 (0.51, 0.81) | 0.7 (0.59, 0.82) | 0.7 (0.59, 0.82) | 0.22 (0, 0.48) | 19 |
| Uzbekistan | EUR | lmi | 0.65 (0.5, 0.8) | 0.7 (0.58, 0.81) | 0.9 (0.79, 1) | 0.81 (0.56, 1) | 19 |
| St. Vincent & the Grenadines | AMR | umi | 0.82 (0.69, 0.95) | 0.87 (0.75, 0.99) | 0.85 (0.77, 0.94) | 0.71 (0.48, 0.95) | 19 (assumption for survival rate of anal, vaginal, and vulvar cancers based on AMR) |
| Venezuela, Bolivarian Republic of | AMR | | 0.66 (0.53, 0.78) | 0.83 (0.71, 0.94) | 0.79 (0.7, 0.87) | 0.76 (0.52, 0.99) | 19 |
| Viet Nam | WPR | lmi | 0.71 (0.6, 0.82) | 0.9 (0.84, 0.97) | 0.94 (0.8, 1) | 0.87 (0.68, 1) | 19 |
| Vanuatu | WPR | lmi | 1 (0.89, 1) * | 1 (0.93, 1) * | 0.52 (0.37, 0.66) | 1 (0.81, 1) * | 19 |
| Samoa | WPR | | 1 (0.89, 1) * | 1 (0.93, 1) * | 1 (0.85, 1) * | 1 (0.81, 1) * | 19 |
| Kosovo | EUR | umi | 0.76 (0.61, 0.91) | 0.76 (0.64, 0.87) | 0.82 (0.7, 0.94) | 0.41 (0.15, 0.66) | 19 (assumption for survival rate of all cancers based on EUR) |

| | | | | | | | |
|---|---|---|---|---|---|---|---|
| Yemen | EMR | low | 0.95 (0.86, 1) | 0.79 (0.74, 0.85) | 0.85 (0.74, 0.97) | 0.93 (0.8, 1) | 19 |
| South Africa | AFR | umi | 0.8 (0.69, 0.92) | 0.84 (0.75, 0.93) | 0.83 (0.74, 0.91) | 0.58 (0.38, 0.77) | 19 |
| Zambia | AFR | lmi | 0.71 (0.6, 0.82) | 0.99 (0.9, 1) | 0.77 (0.68, 0.86) | 0.44 (0.25, 0.63) | 19 |
| Zimbabwe | AFR | lmi | 0.56 (0.45, 0.67) | 0.91 (0.82, 1) | 0.76 (0.67, 0.84) | 0.38 (0.19, 0.57) | 19 |

**Table S8:** Probability of death and uncertainty bounds across countries in analysis.

**Note:** * Indicates where adjustments were made for the survival rate bounds. Calculations for probability of death and its bounds were based on the relative mortality rates (per 100,000 women per year) from the HPV Stats database.[19] AFR= African Region; AMR= Region of the Americas; EMR= Eastern Mediterranean Region; EUR= European Region; lb= lower bound; lmi= lower-middle income; ub= upper bound; umi= upper-middle income; WPR= Western Pacific Region.

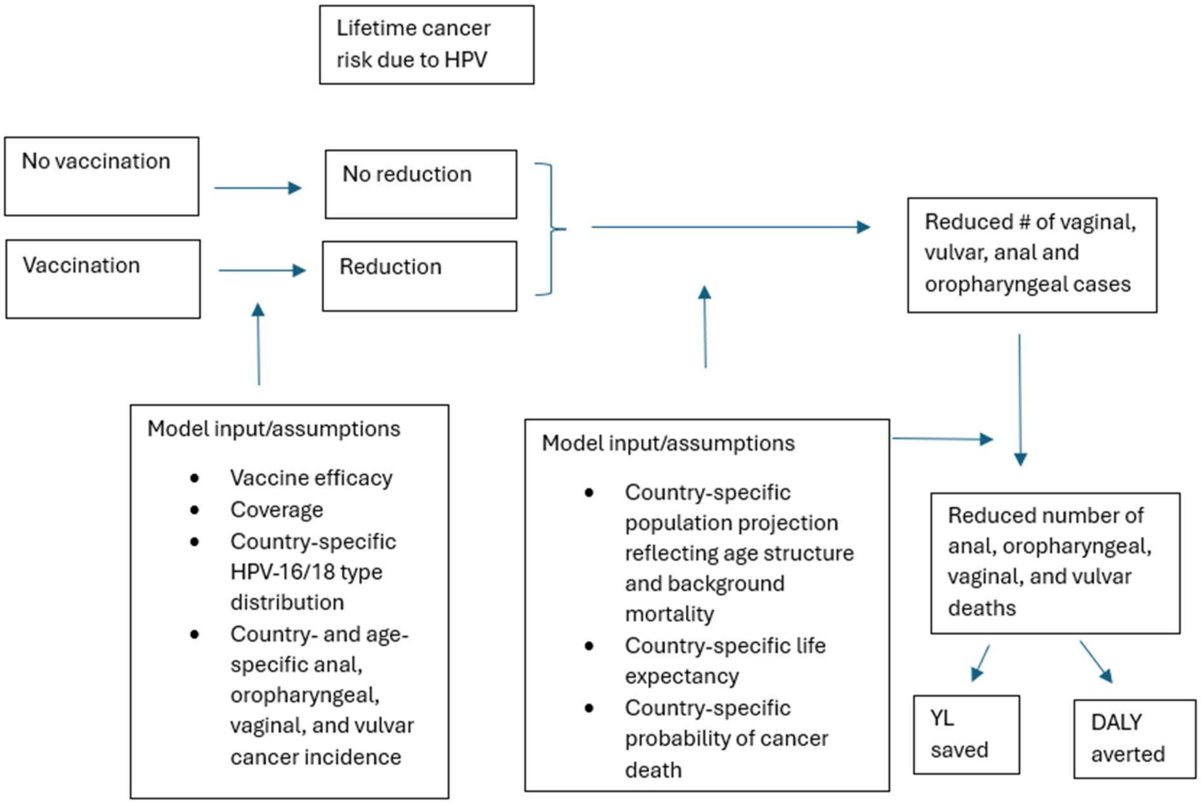

**Figure S1:** An overview of the model framework.

Note: YL refers to Years of Life and DALY refers to Disability-Adjusted Life Years.

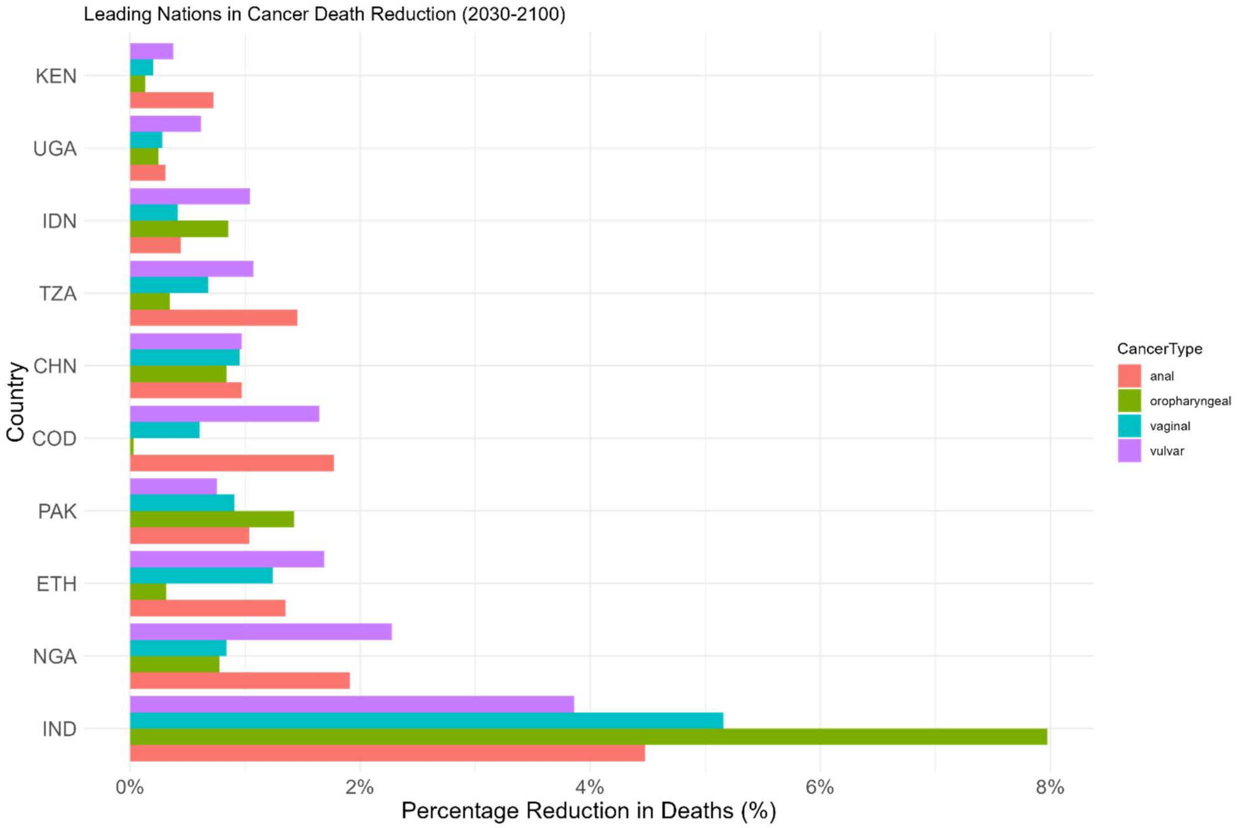

**Figure S2:** Countries leading in deaths averted by HPV vaccination by cancer type.
